# Inequalities in Accident and Emergency department attendance by socio-economic characteristics: population based study

**DOI:** 10.1101/2023.10.10.23296793

**Authors:** Owen Gethings, Perrine Machuel, Vahe Nafilyan

**Affiliations:** Office for National Statistics, Government Buildings, Cardiff Rd, Duffryn, Newport NP10 8XG; Department of Medical Statistics, London School of Hygiene and Tropical Medicine, London, UK

## Abstract

**Objectives:** To examine the relationship between deprivation and Accident and Emergency department attendance.

**Design:** Retrospective cohort study.

**Setting:** England, United Kingdom, from 21 March 2021 to March 2022

**Participants:** All individuals in the 2021 Census, aged 0 to 95 with an Emergency Department attendance record within the Emergency Care Dataset (ECDS). Our full sample included 51,776,958 individuals and 11,498,520 A&E attendance records.

**Main outcome measures:** The primary outcome was any visit to an Accident and Emergency service in England between 21st March 2021 and 31st March 2022 as recorded in ECDS.

**Results:** After adjusting for age, sex and ethnicity, the odds of A&E attendance increased as the level of deprivation increased, with the odds for those in the most deprived decile being 1.69 (95% CI – 1.68 to 1.69) times greater than those in the least deprived decile. Adjusting for underlying health attenuated but did not fully explain the association between deprivation and A&E attendance, with the odds ratio of attendance for those in the most deprived decile reduced to 1.41 (95% CI – 1.40 to 1.41). This pattern was similar across age groups however the gradient of the slope was steeper for working age adults and the magnitude of the reduction in odds for the most deprived decile relative to the least deprived decile after adjusting for health was greatest in those aged 30 to 79. By acuity, those living in the most deprived decile had 2.26 times (95% CI = 2.23 to 2.28) higher odds of attending A&E for a condition classified as low acuity compared with those in the least deprived decile. Even after adjusting for health, those in the most deprived decile had 2.02 (95% CI = 1.99 to 2.02) times the odds of attending for a low acuity condition compared with those in the least deprived decile. This was true for all levels of acuity, except those classified as immediate care, where after adjustment for health, those in the most deprived decile had 0.83 (95% CI = 0.82 to 0.85) times the odds of attendance compared with those in the least deprived decile.

**Conclusions:** People living in more deprived areas were more likely to access A&E services than those living in less deprived areas and these differences are not fully explained by differences in underlying health. The differences were larger for A&E attendance for less severe conditions. Differences in access to primary care services may explain part of these differences in A&E access. Knowing which groups are more likely to attend A&E services will give valuable insight for health services providers, and allow decision makers to better understand how populations can access care differently depending on a range of factors.

**Key messages:** *What is already known on this subject:*

- Previous work has found a clear link between deprivation and health.
- Small-scale or single-centre studies have found links between deprivation and Accident and Emergency attendance.

*What this study adds:*

- This study of 51,776,958 people, and 11,498,520 people with at least one Accident and Emergency department attendance shows a clear deprivation effect, even after adjusting for underlying health.
- People living in more deprived areas were more likely to attend A&E, particularly for low conditions classed as low acuity.
- Underlying health is less important a driver of attendance patterns for people under 30 and is more important a factor for people aged 30 to 65 years of age.

## Introduction

Demand for emergency services in England reached record levels in the Winter 2022/2023, after steadily increasing between 2011 and 2019 [1], with 6 million A&E attendances being recorded between January and March 2023 [2]. The reductions in service provision during the Covid-19 pandemic resulted in a significant backlog for care, which increased pressure on A&E services [3]. The pressure is also due to people attending A&E for conditions that could be treated in primary care or elsewhere in the NHS: A systematic review conducted in 2009 found that between 20 and 40% of A&E attendances are for non-urgent conditions that could be treated elsewhere [4]. Understanding which groups disproportionally use A&E services for non-emergency care could help target interventions.

Socio-economically disadvantaged people are more frequent users of healthcare services in general [5] and also of emergency services, with evidence that those living in the most deprived areas are twice as likely to attend emergency services as those in the least deprived areas [6]. Reasons for A&E attendance are complex and although previous work has shown a clear link between deprivation and A&E attendance [6–9], the underlying cause of this remains unclear. Although the link between deprivation and prevalence of individual chronic diseases [10] and multimorbidity has been well established [11], some evidence suggests that more deprived individuals attend ED because of difficulties accessing General Practices and other community services [12]. Indeed, recent data from England suggests that the number of General Practitioners is lower per head in more deprived communities, despite lower health in these populations [13]. However, to date, there is no study assessing socio-economic differences in A&E attendance using population wide data in England and investigating whether these differences are driven by differences in health.

In this study, we used Census 2021 data linked with emergency care data to examine the socio-economic inequalities in A&E attendances in England between March 2021 and March 2022 and assess whether these differences are driven by differences in health status between the groups.

## Methods

### Study design and data

We used a person-level dataset comprising of individuals in the 2021 Census, linked to the Personal Demographics Service to obtain NHS numbers with a linkage rate of 94.6%. Age was truncated at 95 and these individuals were then linked via NHS number to NHS England’s Emergency Care Data set (ECDS) on Emergency Department attendances. ECDS is the national data set for urgent and emergency care and replaced Accident and Emergency Commissioning Data Set and provides information to support the care provided in emergency departments by including the data items needed to understand capacity and demand and help improve patient care.

The Hospital Episode Statistics (HES) dataset for Admitted Patient Care (APC) and Outpatients (OP) was used to obtain health data to determine patient’s medical history between 01 January 2017 and 21 March 2021. The linked dataset included data on 51,776,958 people in England, which covers approximately 91.7% of the population of England on census day 2021.

### Outcome

The primary outcome was any visit to an A&E service in England between 21st March 2021 and 31^st^ March 2022 as recorded in ECDS. We included all Type 1 General Emergency Departments, Type 2 Specialist Emergency Departments (e.g. paediatric, ophthalmology), Type 3 Minor Injury Units and Type 4 Walk in Centres. As a secondary outcome, we stratified visits by acuity (ECDS codes; 1) Immediate Care, 2) Very Urgent Care, 3) Urgent Care, 4) Standard Care, 5) Low Acuity).

### Exposures

Our main exposure of interest was Socio-Economic Status (SES), as measured by deciles of the Index of Multiple Deprivation 2019 (IMD) of the area of residence at the time of the 2021 Census. As SES is a complex concept we used several different measures as alternative in our analyses, which are presented in supplementary materials. We used information from the 2021 census on Level of highest qualification (“No qualifications”, “Apprenticeship”, “A levels, AS levels and equivalent”, “5+ GCSEs and equivalent”, “1-4 GCSEs and equivalent”, “Degree and above”) and individual National Statistics Socio-economic classification (NS-SEC) (“Never worked and long term unemployed”, “Routine occupations”, “Semi-routine occupations”, “Lower supervisory and technical occupations”, “Small employers and own account workers”, “Intermediate occupations”, “Lower managerial, administrative and professional occupations” and “Higher managerial, administrative and professional occupations”).

### Covariates

To estimate the differences in access by SES status, we fitted models that were adjusted for a range of confounding factors. Age was included as a restricted cubic spline with knots at the 1^st^, 50^th^ and 99^th^ percentile. We also included sex (Male, Female), and ethnicity (White, Asian, Black, Mixed/Multiple Ethnic Group, Other).

To assess whether the differences in access by SES were driven by differences in health, we fitted models that were also adjusted for individual health. Prevalence of health conditions was measured using data from HES APC/OP records (see supplementary table 1). We also included measures of health collected at the 2021 Census, including long-term health or disability (“Not Limited”, “Yes – reduced a little”, “Yes – reduced a lot”, “Yes – not reduced at all”), and self-reported general health (“Very Good Health”, “Good Health”, “Fair Health”, “Bad Health”) as well as using information from HES APC records (List conditions included). To proxy severity, we also created a variable for the number of provider spells from HES APC in the years prior to our study period, covering the period 1^st^ January 2017 to 21^st^ March 2021.

### Statistical analysis

Characteristics of the study population were summarised overall, using means for continuous variables and proportions for categorical variables. We calculated absolute risk by age and sex by estimating the proportion of people who attended A&E by sex and single year of age.

To estimate the differences in access by SES status, we fitted Generalized Linear Models with binomial errors and a logit link to estimate the odds of A&E attendance whilst controlling for patient characteristics as detailed above. First, we examined the differences in emergency attendance by SES, by fitting models adjusted for confounding factors only (age, sex, ethnicity). Second, we assessed to what extent health mediated the relationship between our exposures and emergency attendance by adjusting for health variables. To do so we included measures of health to the models and compared the odds ratios of the health-adjusted model to the odds ratios of the confounder-adjusted model. We included interactions between age and sex, age and ethnicity, age and disability, age and health in general, age and provider spell count and age and all HES health variables. We also included interactions between sex and ethnicity, sex and disability, sex and health in general, sex and provider spell count and sex and all HES health variables to ensure that confounder factors were appropriately adjusted for. Due to collinearity between socio-economic variables, we fitted separate regression models for each exposure.

Analyses were then further stratified by age group (0-5, 6-15, 16-29, 30-49, 50-64, 65-79 and 80 to 95) and acuity. Models with NSSEC and Level of highest Qualification as exposures were restricted to residents aged 25+ (25-49, 50-64, 65-79 and 80 to 95). Age was included as a restricted cubic spline. We included interactions for sex and ethnicity, sex and disability, sex and health in general, sex and provider spell count and sex and all HES health variables. Relative reduction in odds between the unadjusted and health-adjusted models was calculated by *(OR_1 – OR_2)/(OR_2-1)*, where *OR_1* is the health-adjusted model odds ratio and *OR_2* is the confounder-adjusted model odds ratio.

### Patient and public involvement

We did not directly involve patients and the public in the design and conception of the study, primarily because this study was conducted rapidly.

## Results

### Characteristics of the study population

Our full individual-level dataset set contained 51,776,958 individuals. The average age was 41 (±23.7), and 48% were male (Table 1).

**Table 1.** Characteristics of the full study population, and characteristics of population with at least one Accident and Emergency attendance.

| Variable | Levels | Characteristics of the full study population |  | Attended A&E |  |
| --- | --- | --- | --- | --- | --- |
|  |  | Number | Percent | Number | Percent |
| Sex | Male (Reference) | 24,939,885 | 48.2% | 5,451,800 | 47.4% |
| Sex | Female | 26,837,075 | 51.8% | 6,046,720 | 52.6% |
| Age group | <5 | 3,465,435 | 6.7% | 1,183,370 | 10.3% |
| Age group | 6 to 17 years old | 7,575,290 | 14.6% | 1,630,645 | 14.2% |
| Age group | 18 to 29 years old | 6,968,920 | 13.5% | 1,659,520 | 14.4% |
| Age group | 30 to 49 years old | 13,481,765 | 26.0% | 2,605,620 | 22.7% |
| Age group | 50 to 64 years old | 10,371,340 | 20.0% | 1,914,680 | 16.7% |
| Age group | 65 to 79 years old | 7,332,785 | 14.2% | 1,567,305 | 13.6% |
| Age group | 80 to 95 years old | 2,581,425 | 5.0% | 937,385 | 8.2% |
| Ethnicity | White (Reference) | 42,447,695 | 82.0% | 9,369,880 | 81.5% |
| Ethnicity | Black | 1,990,730 | 3.8% | 477,400 | 4.2% |
| Ethnicity | Mixed/multiple ethnic group | 1,472,185 | 2.8% | 358,840 | 3.1% |
| Ethnicity | Asian | 4,804,180 | 9.3% | 1,030,515 | 9.0% |
| Ethnicity | Other | 1,062,165 | 2.1% | 261,885 | 2.3% |
| IMD Decile | 1 | 5,017,900 | 9.7% | 1,369,020 | 11.9% |
| IMD Decile | 2 | 5,105,645 | 9.9% | 1,303,325 | 11.3% |
| IMD Decile | 3 | 5,191,665 | 10.0% | 1,253,000 | 10.9% |
| IMD Decile | 4 | 5,198,220 | 10.0% | 1,195,605 | 10.4% |
| IMD Decile | 5 | 5,206,780 | 10.1% | 1,140,645 | 9.9% |
| IMD Decile | 6 | 5,289,110 | 10.2% | 1,121,790 | 9.8% |
| IMD Decile | 7 | 5,195,920 | 10.0% | 1,074,875 | 9.3% |
| IMD Decile | 8 | 5,227,770 | 10.1% | 1,062,295 | 9.2% |
| IMD Decile | 9 | 5,186,325 | 10.0% | 1,019,975 | 8.9% |
| IMD Decile | 10 (Reference) | 5,157,620 | 10.0% | 957,995 | 8.3% |
| Health in general | Very good health (Reference) | 24,938,600 | 48.2% | 4,806,405 | 41.8% |
| Health in general | Good health | 17,523,130 | 33.8% | 3,653,440 | 31.8% |
| Health in general | Fair health | 6,627,135 | 12.8% | 1,952,965 | 17.0% |
| Health in general | Bad health | 2,082,770 | 4.0% | 815,890 | 7.1% |
| Health in general | Very bad health | 605,320 | 1.2% | 269,820 | 2.3% |
| Disability | Not limited (Reference) | 39,089,090 | 75.5% | 7,775,155 | 67.6% |
| Disability | Yes - not reduced at all | 3,626,320 | 7.0% | 788,235 | 6.9% |
| Disability | Yes - reduced a little | 5,249,275 | 10.1% | 1,517,625 | 13.2% |
| Disability | Yes - reduced a lot | 3,812,270 | 7.4% | 1,417,505 | 12.3% |
| The National Statistics Socio-economic classification | Higher managerial administrative and professional occupations (Reference) | 5,635,755 | 10.9% | 895,330 | 7.8% |
| The National Statistics Socio-economic classification | Lower managerial, administrative and professional occupations | 8,494,045 | 16.4% | 1,607,350 | 14.0% |
| The National Statistics Socio-economic classification | Intermediate occupations | 4,915,500 | 9.5% | 1,021,730 | 8.9% |
| The National Statistics Socio-economic classification | Small employers and own account workers | 4,450,280 | 8.6% | 936,360 | 8.1% |
| The National Statistics Socio-economic classification | Lower supervisory and technical occupations | 2,248,500 | 4.3% | 514,190 | 4.5% |
| The National Statistics Socio-economic classification | Semi-routine occupations | 4,825,120 | 9.3% | 1,154,490 | 10.0% |
| The National Statistics Socio-economic classification | Routine occupations | 5,060,880 | 9.8% | 1,226,295 | 10.7% |
| The National Statistics Socio-economic classification | Never worked and long-term unemployed | 3,499,995 | 6.8% | 970,120 | 8.4% |
| The National Statistics Socio-economic classification | Not classified | 12,646,885 | 24.4% | 3,172,655 | 27.6% |
| Level of highest qualification | No qualifications | 7,601,715 | 14.7% | 2,016,425 | 17.5% |
| Level of highest qualification | Other or not classified | 11,008,585 | 21.3% | 2,827,375 | 24.6% |
| Level of highest qualification | Apprenticeship | 2,264,315 | 4.4% | 537,480 | 4.7% |
| Level of highest qualification | Level 1: 1-4 GCSEs and equivalent | 4,130,020 | 8.0% | 922,610 | 8.0% |
| Level of highest qualification | Level 2: 5+ GCSEs and equivalent | 5,659,000 | 10.9% | 1,213,940 | 10.6% |
| Level of highest qualification | Level 3: A levels, AS levels and equivalent | 6,871,590 | 13.3% | 1,425,140 | 12.4% |
| Level of highest qualification | Degree level and above (Reference) | 14,241,735 | 27.5% | 2,555,555 | 22.2% |

Between 21st March 2021 and 31^st^ March 2022, 11,498,520 (22%) people attended an A&E service in England at least once (Mean number of visits = 1.7 ± 1.6 sd). Out of all attendances, 1,369,021 (12%) were among those from the most deprived decile, with 957,993 (8%) being from the least-deprived decile. 53% of attendances were among females.

### A&E attendance by age and sex

Figure 1 shows the proportion of people having attended A&E at least once between 21st March 2021 and 31^st^ March 2022 by age and sex. The proportion of people who attended an emergency department was highest in infants (aged 0 to 2 years old) and in older adults (aged 80 years old and above). The proportion of people having attended A&E generally decreased with age from ages 2 to 10, with increases for males between the ages of 10 and 20, and an increase for women between the ages of 17 and 29. It remained low for both sexes until around the age of 70 where it then increased with age.

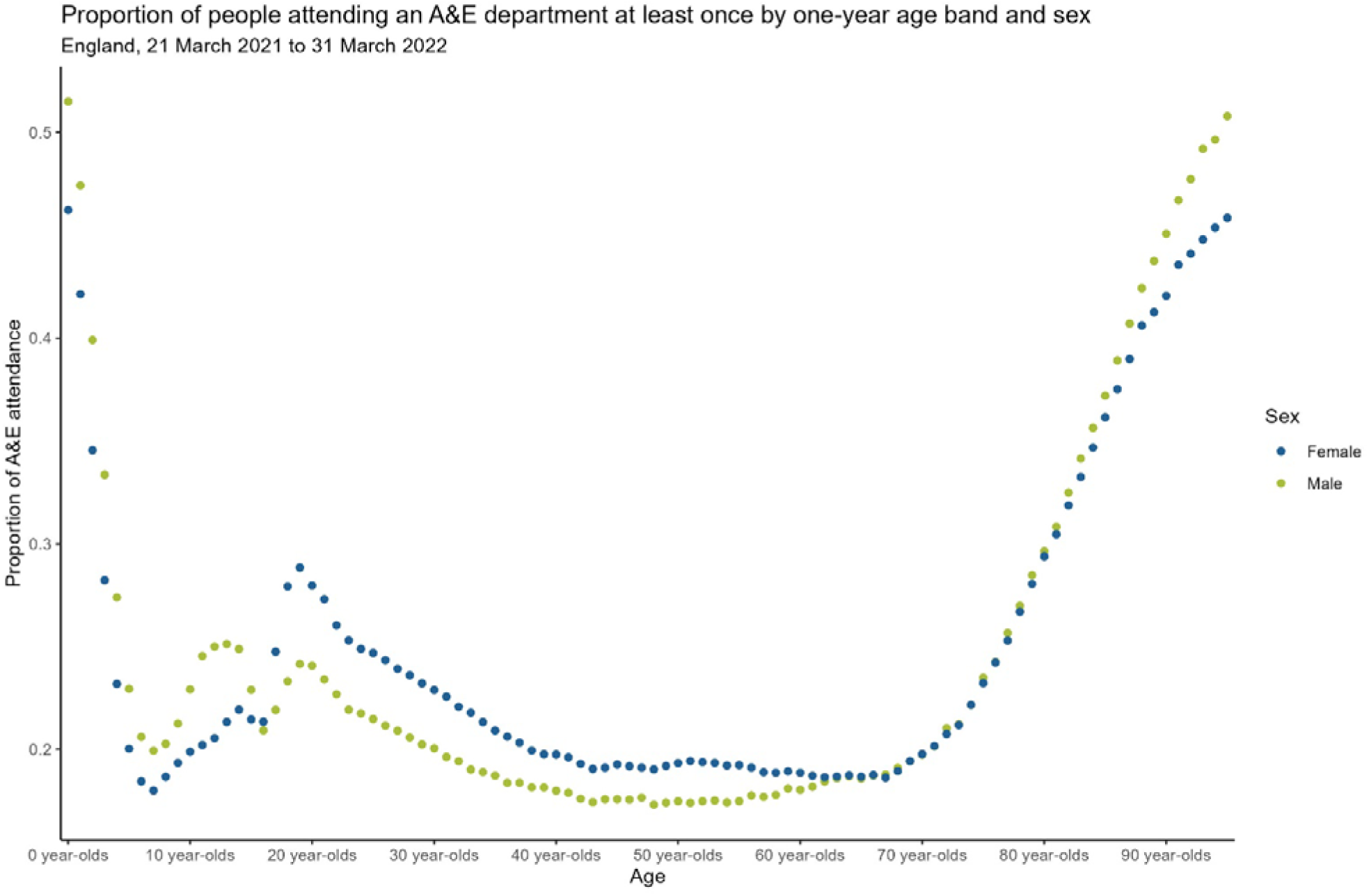

#### Index of Multiple Deprivation

Odds ratios by IMD decile are given in table 2. After adjusting for age, sex and ethnicity, the odds of A&E attendance increased as the level of deprivation increased, with the odds for those in the most deprived decile being 1.69 (95% CI – 1.68 to 1.69) times greater than for those in the least deprived decile. After adjusting for underlying health, overall, the odds of A&E attendance again increased as deprivation increased, however, the odds of attendance for those in the most deprived decile reduced to 1.41 (95% CI – 1.40 to 1.41). This corresponds to a 40.9% (95% CI – 40.7% to 41.1%) difference in odds after adjusting for underlying health.

**Table 2.** Odds ratios for Accident and Emergency Attendance by IMD for unadjusted and health-adjusted models.

| Level | Model adjustment | Odds ratio | lower 95% | Upper 95% |
| --- | --- | --- | --- | --- |
| IMD Decile 1 | Health adjusted | 1.41 | 1.40 | 1.41 |
| IMD Decile 1 | Not health adjusted | 1.69 | 1.68 | 1.69 |
| IMD Decile 2 | Health adjusted | 1.34 | 1.33 | 1.34 |
| IMD Decile 2 | Not health adjusted | 1.55 | 1.54 | 1.55 |
| IMD Decile 3 | Health adjusted | 1.27 | 1.27 | 1.28 |
| IMD Decile 3 | Not health adjusted | 1.44 | 1.44 | 1.44 |
| IMD Decile 4 | Health adjusted | 1.22 | 1.21 | 1.22 |
| IMD Decile 4 | Not health adjusted | 1.35 | 1.34 | 1.35 |
| IMD Decile 5 | Health adjusted | 1.16 | 1.16 | 1.16 |
| IMD Decile 5 | Not health adjusted | 1.26 | 1.25 | 1.26 |
| IMD Decile 6 | Health adjusted | 1.13 | 1.12 | 1.13 |
| IMD Decile 6 | Not health adjusted | 1.20 | 1.20 | 1.20 |
| IMD Decile 7 | Health adjusted | 1.10 | 1.10 | 1.11 |
| IMD Decile 7 | Not health adjusted | 1.16 | 1.15 | 1.16 |
| IMD Decile 8 | Health adjusted | 1.09 | 1.09 | 1.09 |
| IMD Decile 8 | Not health adjusted | 1.13 | 1.12 | 1.13 |
| IMD Decile 9 | Health adjusted | 1.06 | 1.05 | 1.06 |
| IMD Decile 9 | Not health adjusted | 1.08 | 1.08 | 1.08 |

Figure 2 shows A&E attendance by age and IMD decile.

Odds ratios for deprivation by age are given in table 3 for decile 1, other deciles are given in supplementary table 2. Across all age groups, the odds of attendance increased as the level of deprivation increased, but the gradient was steeper for working age adults than for older adults and children. Prior to adjusting for health, 30 to 49 (OR = 1.86 95% CI - 1.85 to 1.87), 50 to 64 (OR = 1.87, 95% CI - 1.86 to 1.88) and 65 to 79 years olds (OR = 1.73, 95% CI – 1.71 to 1.74) living in the 10% most deprived areas (decile 1) had the greatest odds of A&E attendance, relative to their counterparts in decile 10. After adjusting for health, we observed a substantial reduction in the odds of A&E attendance. The odds of attendance for those in decile 1 compared with decile 10 reduced substantially for 30 to 49 (OR = 1.54, 95% CI – 1.53 to 1.55), 40 to 64 (OR = 1.39, 95% CI – 1.38 to 1.40) and 65 to 79-year-olds (OR = 1.23, 95% CI – 1.22 to 1.24) In relative terms (Table 3), zero to 5-year-olds had the smallest relative reduction in odds (10.7% 95% CI – 10.5% to 11.1%) after adjusting for health, with 65 to 79-year-olds seeing the largest reduction (68.5%, 67.6% to 69.0%).

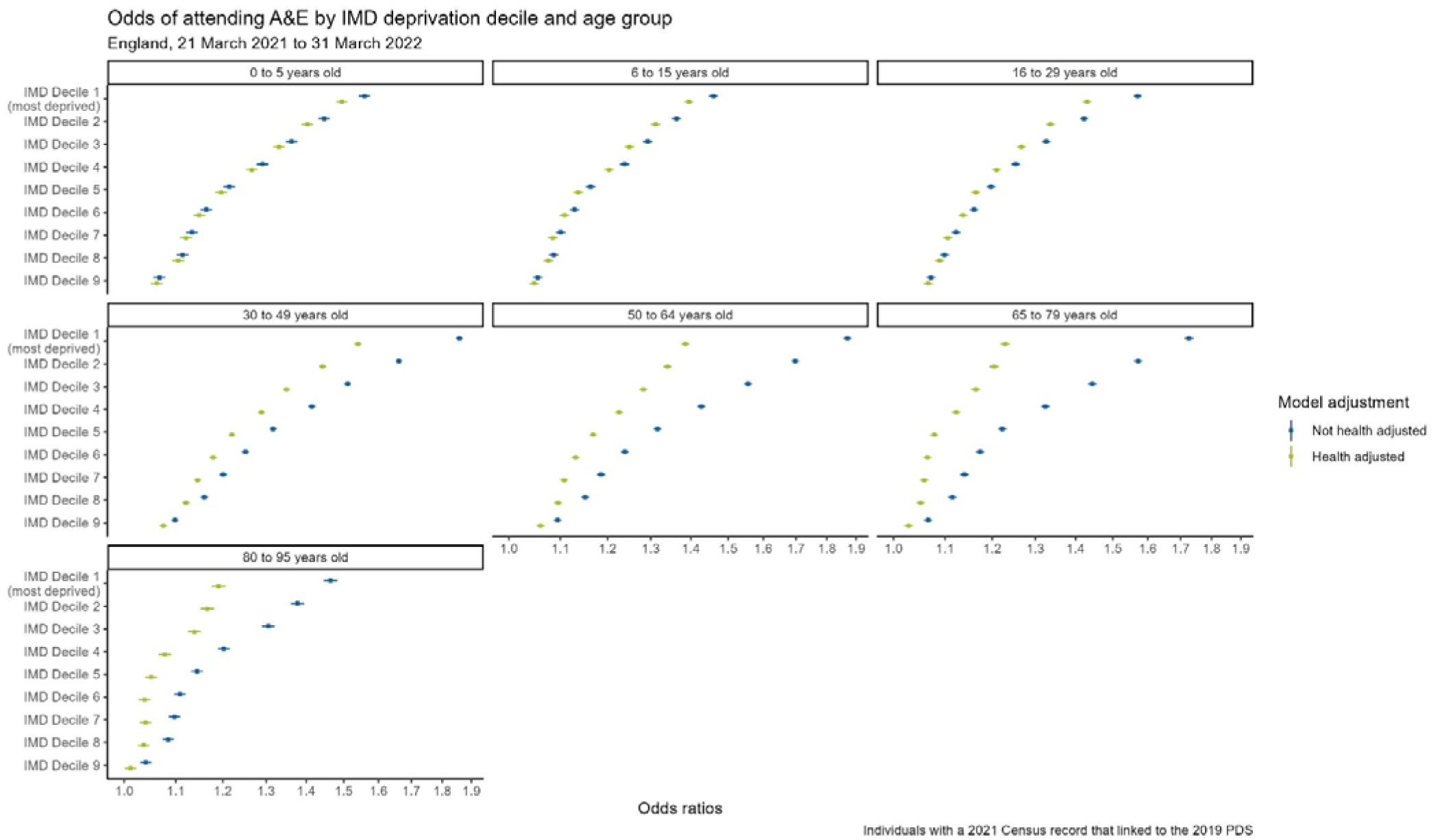

**Table 3.** Odds ratios by age and IMD for unadjusted and health-adjusted models with lower and upper 95% confidence intervals. Percent relative reduction between unadjusted and health-adjusted models.

| IMD Decile | Model type | Age group | Odds ratio | lower 95% | Upper 95% | Relative reduction | Lower 95% Relative reduction | Upper 95% relative reduction |
| --- | --- | --- | --- | --- | --- | --- | --- | --- |
| IMD Decile 1 | Health adjusted | 0 to 5 years old | 1.50 | 1.48 | 1.51 | -10.7% | -11.1% | -10.5% |
| IMD Decile 1 | Not health adjusted | 0 to 5 years old | 1.56 | 1.54 | 1.57 |  |  |  |
| IMD Decile 1 | Health adjusted | 6 to 15 years old | 1.40 | 1.38 | 1.41 | -13.0% | -15.6% | -12.8% |
| IMD Decile 1 | Not health adjusted | 6 to 15 years old | 1.46 | 1.45 | 1.47 |  |  |  |
| IMD Decile 1 | Health adjusted | 16 to 29 years old | 1.43 | 1.42 | 1.44 | -24.6% | -25.0% | -24.1% |
| IMD Decile 1 | Not health adjusted | 16 to 29 years old | 1.57 | 1.56 | 1.58 |  |  |  |
| IMD Decile 1 | Health adjusted | 30 to 49 years old | 1.54 | 1.53 | 1.55 | -37.2% | -37.6% | -36.8% |
| IMD Decile 1 | Not health adjusted | 30 to 49 years old | 1.86 | 1.85 | 1.87 |  |  |  |
| IMD Decile 1 | Health adjusted | 50 to 64 years old | 1.39 | 1.38 | 1.40 | -55.2% | -55.8% | -54.5% |
| IMD Decile 1 | Not health adjusted | 50 to 64 years old | 1.87 | 1.86 | 1.88 |  |  |  |
| IMD Decile 1 | Health adjusted | 65 to 79 years old | 1.23 | 1.22 | 1.24 | -68.5% | -69.0% | -67.6% |
| IMD Decile 1 | Not health adjusted | 65 to 79 years old | 1.73 | 1.71 | 1.74 |  |  |  |
| IMD Decile 1 | Health adjusted | 80 to 95 years old | 1.19 | 1.18 | 1.21 | -58.7% | -60.0% | -56.3% |
| IMD Decile 1 | Not health adjusted | 80 to 95 years old | 1.46 | 1.45 | 1.48 |  |  |  |

Figure 3 shows A&E attendance by acuity and IMD decile. Odds ratios of A&E attendance for different levels of acuity by deprivation are given in table 4 for decile 1, other deciles are given in supplementary table 3. For low acuity (5), standard care (4), urgent care (3) and very urgent care (2), the odds of attendance increased as deprivation increased. The relationship was less clear for immediate care (1), with those in decile 3 (OR = 1.46, 95% CI = 1.44 to 1.49) and 5 (OR = 1.43, 95% CI = 1.40 to 1.46) having the highest odds of attendance. Those living in the most deprived decile had 2.26 times (95% CI = 2.23 to 2.28) higher odds of attending A&E for a condition classified as low acuity compared with those in the least deprived decile. After adjusting for underlying health, again, for low acuity (5), standard care (4), urgent care (3) and very urgent care (2), the odds of attendance increased as deprivation increased. Those in the most deprived decile had 2.02 (95% CI = 1.99 to 2.02) times the odds of attending for a low acuity condition compared with those in the least deprived decile. However, for immediate level care, odds of attendance generally decreased as deprivation increased, with those in the most deprived decile having 0.83 (95% CI = 0.82 to 0.85) times the odds of attendance compared with those in the least deprived decile. Those in the most deprived decile had 2.02 (95% CI = 1.99 to 2.02) times the odds of attending for a low acuity condition compared with those in the least deprived decile.

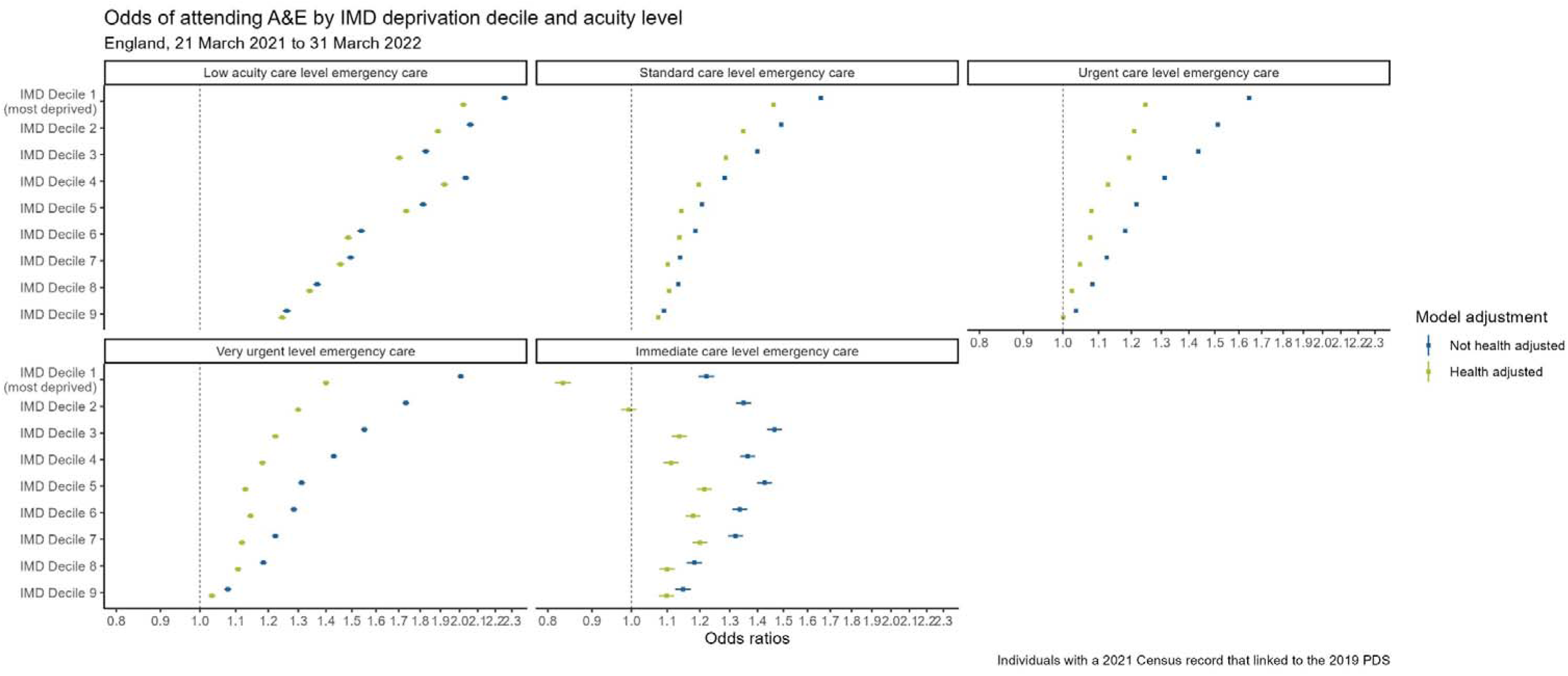

**Table 4.**
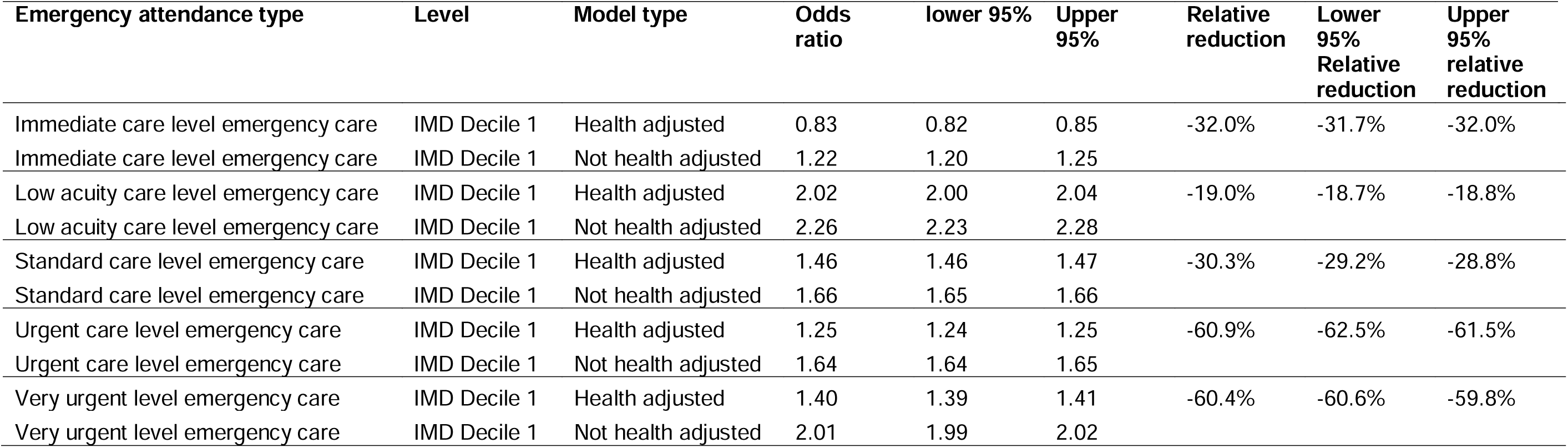
Odds ratios by acuity and IMD for unadjusted and health-adjusted models with lower and upper 95% confidence intervals. Percent relative reduction between unadjusted and health-adjusted models.

Standard Care (30.3%, 28.8% to 29.2%), Urgent Care (60.9%, 61.5% to 62.5%), Very Urgent Care (60.4%, 59.8% to 60.6%) and Immediate Care (32.0%, 31.7% to 32.0%) saw the largest relative reduction in odds between health-unadjusted and adjusted models, with Low Acuity seeing only a 19.0% (18.7% to 18.8%) relative reduction.

Acuity by age

#### Other measures of Socioeconomic status

Results for our other proxy measures of socioeconomic status can be found in supplementary table 2 and 3. Patterns and results were similar for other SES positions.

## Discussion

### Main findings

In this study using a unique linkage of the 2021 Census with emergency care data, we found people living in more deprived areas were more likely to have attended A&E at least once, after adjusting for age, sex and ethnicity. This difference in A&E access by area deprivation was observed for all age groups but was most pronounced for people aged between 30 and 65 and decreased for older ages.

Adjusting for health using comorbidities data from HES, self-reported general health, self-reported disability status and number of hospital admissions in the 4 years prior to our study partly, but not totally, explained the differences in attendance patterns. Adjusting for health reduced the odds ratios of accessing A&E between the most and least deprived area by about 40%. This pattern was consistent across all age groups, however the relative reduction in odds was greatest for patients aged 30 to 79, with health adjustments having very little effect for people less than 30.

We observed greater socio-economic differences in A&E attendance for low acuity conditions than for higher acuity conditions. The odds of attending A&E for a low acuity condition were 2 times greater for people living in the 10% most deprived areas, relative to their counterparts in the 10% least deprived areas. Further adjusting for health reduced the estimated differences in A&E attendance but the overall pattern remained the same: people in the most deprived deciles had greater odds of attendance for low acuity conditions than for high acuity conditions. These heightened differences in service use for conditions of low acuity for people in the most deprived areas cannot be explained by differences in age, sex, ethnicity, or underlying health.

### Comparison with other studies

Consistent with our findings, previous work has found a clear relationship between deprivation and A&E attendances [6,7,8,13,15,16], with people from more deprived backgrounds generally having higher odds of attending an A&E service in England. Most of these previous studies have either been based on small national surveys [6, 14], restricted to specific geographies [7,8,9,16] which may not reflect patterns observed elsewhere or did not adjust for underlying health [6, 7, 15]. A large analysis of 17 million attendances by LSOA extracted from HES between April 2011 and March 2012 found that the number of both inappropriate and appropriate attendances increased as deprivation increased [15]. However, they also found that after controlling for age, sex and gender, those from the least deprived quintile had the greatest odds of inappropriate attendances relative to appropriate attendances. In contrast to these previous studies, [9] found that IMD was not a predictor for A&E attendance, however they specifically only chose disadvantaged neighbourhoods, where the variation in IMD between areas was perhaps not sufficient to detect a meaningful association.

Many of these studies have hypothesised that the differences in attendance rates by deprivation are largely driven by differences in underlying health. Indeed, the link between deprivation and prevalence of individual chronic diseases [10] and multimorbidity has been well established [11]. However, we find that even after adjusting for health, using a comprehensive set of measures of underlying health, people living in more deprived areas are more likely to attend A&E than those living in less deprived areas.

After adjusting for health, we found the difference between the most and least deprived 10% of areas decreased above the age of 65. This suggests that deprivation is less important a factor than general health, age, or perhaps frailty. Indeed, relative differences between the unadjusted and health-adjusted odds ratios were greatest for those 30 to 49, 50 to 64 and 65 to 79 years, with the biggest relative reduction seen in those aged 65 to 79 years of age. This is consistent with findings from an analysis of NHS New Devon CCG data [16] and Glasgow A&E resident data [18], that found that over age 65, the biological effects of ageing outweigh the social effects of deprivation [16].

Very few studies have assessed differences in attendance by clinical acuity across the deprivation gradient. There is evidence that people from more deprived backgrounds were more likely to be classed as “more severe” upon arrival at A&E [19]. However, they defined severity based on a subjective analysis of diagnosis codes, limited their sample to ambulance arrivals and excluded self-referrals. We used a standardised measure of acuity available within ECDS to limit subjectivity and ensure findings were applicable across areas. Another study looking at children under the age of 13 found that there was a trend for an increased attendance in all triage categories for the most deprived populations [20]. We found increased odds of attendance for people in the most deprived deciles for all acuity classifications except immediate level care. An analysis from Canada also found that materially deprived residents, particularly young adults, used the emergency department disproportionately more than the least deprived do, generally for all medical conditions and particularly for low-acuity conditions [21]. This was especially true for low acuity conditions during the COVID-19 pandemic [22]. These heightened differences in service use for conditions of low acuity for people in the most deprived areas cannot be explained solely by differences in underlying health but could be driven by other factors such as access to primary care [12, 23, 24].

The reason for greater attendance rates, particularly for low acuity conditions, among people from lower SES is currently unclear. Previous work has found that young people from lower SES are more likely to attend A&E for injuries [25] and unintentional poisonings [26], however we did not look at cause-specific attendances within this analysis. Other work has suggested that difficulties accessing primary care services could also play a role [14], as well as dissatisfaction with primary care services [23, 24] and lower General Practice availability among more deprived communities [13]. Recent work has found fewer total GPs, Direct Patient Care staff and paramedics per 10,000 patients in deprived areas [27], with the inequalities widening significantly over time. Similarly, GP practices with high GP turnover have been shown to be significantly associated with more emergency hospital attendances per 100 patients, with turnover being higher in more deprived areas [28]. This could disproportionately affect more deprived patients, who tend to be in poorer health, and attend A&E due to inability to access a GP practice.

### Strengths and limitations of this study

Our study has several strengths. The first, we used a unique population-level dataset based on the 2021 Census. Census 2021 covered around 97% of the population, and therefore is the most representative data source available to produce statistics about the population living in England. Our study is the largest study to examine the association between socio-economic status and A&E attendance to date, and make use of the information provided by Census such as socioeconomic classification and self-reported ethnicity, self-reported general health and disability status.

Second, an important strength of our study is that we combined multiple sources of health data to measure people’s health status at the beginning of the study period. We have objective information on hospital admission from the Hospital Episode Statistics (HES) and, crucially, self-reported general health and disability status from the 2021 Census, which provide richer information on people’s health than what is typically available in studies solely based on electronic health records and enables us to assess whether the differences in A&E access by socio-economic status are driven by differences in health status.

Our study also has some limitations. Not all people living in England in March 2021 were enumerated at Census 2021, and of those who were, not all could be linked to an NHS number via the Personal Demographics Service. Linking to Census invariably means we may exclude some people, for example people without a fixed address who may be living in extremely disadvantaged circumstances were probably not captured by our data. However, our dataset covers 91.7% of the population living in England at the time of Census.

Another limitation is that we only looked at A&E attendances as a binary measure (e.g., whether someone attended an emergency department at least once between 2021 and 2022), which does not discriminate between people who attended A&E only once, and those who attended multiple times. Therefore, we may not be capturing potential compounding effects of SES on the likelihood of attending A&E multiple times.

## Conclusion

Our results suggest that the socio-economic differences in A&E are not solely driven by differences in health, but that other factors such as access to primary care services may explain a large part of these differences in A&E access. Increasing access to primary care services in the most deprived areas could help alleviate the pressure faced emergency departments.

## Ethical approval

This study was ethically self-assessed against the ethical principles of the National Statistician’s Data Ethics Advisory Committee (NSDEC) using NSDEC’s ethics self-assessment tool. We engaged with the UK Statistics Authority Data Ethics team, who were satisfied that no further ethical approval was required.

## Footnotes

Contributors: OG, PM and VN all contributed equally. OG, PM and VN conceived the study. PM performed the methodology, analysis and produced figures. OG, PM and VN interpreted the data. OG and VN drafted the manuscript. All authors revised the manuscript for important intellectual content and approved the final submitted version. OG and PM accessed and verified the data. All authors had full access to the data in the study, acted as guarantors for the study, and had final responsibility for the decision to submit for publication. All authors vouch for the completeness and accuracy of the data and for the fidelity of the trial to the protocol. The corresponding author attests that all listed authors meet authorship criteria and that no others meeting the criteria have been omitted.

## Data Availability

All data produced in the present study are available upon reasonable request to the authors.

## Acknowledgements

We would like to acknowledge NHS England, particularly Mark Svenson for helpful comments about the study design.

## References

[1] Summary Reports - High level - NHS Digital (2021). Available at: https://digital.nhs.uk/data-and-information/publications/statistical/hospital-accident--emergency-activity/2020-21/summary-reports.

[2] A&E waiting times - Nuffield Trust (2023). Available at: https://www.nuffieldtrust.org.uk/resource/a-e-waiting-times

[3] Coronini-Cronberg S, Maile EJ, Majeed A. Health inequalities: the hidden cost of COVID-19 in NHS hospital trusts? Journal of the Royal Society of Medicine. 2020;113(5):179–184. doi:10.1177/0141076820925230

[4] Carret ML, Fassa AC, Domingues MR. Inappropriate use of emergency services: a systematic review of prevalence and associated factors. Cad Saude Publica. 2009 Jan;25(1):7–28. doi: 10.1590/s0102-311×2009000100002. PMID: 19180283.

[5] Saini P, McIntyre J, Corcoran R, Daras K, Giebel C, Fuller E, Shelton J, Wilson T, Comerford T, Nathan R, Gabbay M. Predictors of emergency department and GP use among patients with mental health conditions: a public health survey. Br J Gen Pract. 2019 Dec 26;70(690):e1–e8. doi: 10.3399/bjgp19X707093. PMID: 31848197; PMCID: PMC6917360.

[6] Scantlebury R, Rowlands G, Durbaba S, et al.. Socioeconomic deprivation and accident and emergency attendances: cross-sectional analysis of general practices in England. Br J Gen Pract 2015;65:e649–e654. 10.3399/bjgp15X686893

[7] Blatchford, O., Capewell, S., Murray, S., & Blatchford, M. (1999). Emergency medical admissions in Glasgow: general practices vary despite adjustment for age, sex, and deprivation. British Journal of General Practice, 49(444), 551–554.

[8] Hull, S. A., Homer, K., Boomla, K., Robson, J., & Ashworth, M. (2018). Population and patient factors affecting emergency department attendance in London: retrospective cohort analysis of linked primary and secondary care records. British Journal of General Practice, 68(668), e157–e167.

[9] Giebel C, McIntyre JC, Daras K, et al. What are the social predictors of accident and emergency attendance in disadvantaged neighbourhoods? Results from a cross-sectional household health survey in the north west of England. BMJ Open 2019;9:e022820. doi: 10.1136/bmjopen-2018-022820

[10] Eachus J, Williams M, Chan P, Smith G D, Grainge M, Donovan J et al. Deprivation and cause specific morbidity: evidence from the Somerset and Avon survey of health BMJ 1996; 312:287 doi:10.1136/bmj.312.7026.287

[11] Barnett, K., Mercer, S. W., Norbury, M., Watt, G., Wyke, S., & Guthrie, B. (2012). Epidemiology of multimorbidity and implications for health care, research, and medical education: a cross-sectional study. The Lancet, 380(9836), 37–43.

[12] MacKichan, F., Brangan, E., Wye, L., Checkland, K., Lasserson, D., Huntley, A.,… & Purdy, S. (2017). Why do patients seek primary medical care in emergency departments? An ethnographic exploration of access to general practice. BMJ open, 7(4), e013816.

[13] Fisher R, Dunn P, Asaria M, Thorlby R. Level or not?. The Health Foundation; 2020 (10.37829/HF-2020-RC13).

[14] Shah, S. M., & Cook, D. G. (2008). Socio-economic determinants of casualty and NHS Direct use. Journal of Public Health, 30(1), 75–81.

[15] McHale, P., Wood, S., Hughes, K., Bellis, M. A., Demnitz, U., & Wyke, S. (2013). Who uses emergency departments inappropriately and when-a national cross-sectional study using a monitoring data system. BMC medicine, 11, 1–9.

[16] Pereira Gray D, Henley W, Chenore T, et al. 2017. What is the relationship between age and deprivation in influencing emergency hospital admissions? A model using data from a defined, comprehensive, all-age cohort in East Devon, UKBMJ Open 2017;7:e014045. doi: 10.1136/bmjopen-2016-014045

[17] Johnson, L., Cornish, R., Boyd, A. et al. Socio-demographic patterns in hospital admissions and accident and emergency attendances among young people using linkage to NHS Hospital Episode Statistics: results from the Avon Longitudinal Study of Parents and Children. BMC Health Serv Res 19, 134 (2019). 10.1186/s12913-019-3922-7

[18] K Levin, E Crighton, Sex, age and socioeconomic inequalities in older people’s unscheduled care, European Journal of Public Health, Volume 29, Issue Supplement_4, November 2019, ckz185.142, 10.1093/eurpub/ckz185.142

[19] Turner, A. J., Francetic, I., Watkinson, R., Gillibrand, S., & Sutton, M. (2022). Socioeconomic inequality in access to timely and appropriate care in emergency departments. Journal of Health Economics, 85, 102668.

[20] Beattie, T. F., Gorman, D. R., & Walker, J. J. (2001). The association between deprivation levels, attendance rate and triage category of children attending a children’s accident and emergency department. Emergency Medicine Journal, 18(2), 110–111.

[21] VanStone, N. A., Belanger, P., Moore, K., & Caudle, J. M. (2014). Socioeconomic composition of low-acuity emergency department users in Ontario. Canadian Family Physician, 60(4), 355–362.

[22] Hanscom, D., & Dutton, D. J. (2022). Effect of the COVID-19 pandemic on the socioeconomic composition of emergency department presentations. Canadian Journal of Public Health, 113(6), 878–886.

[23] Agarwal S, Banerjee J, Baker R, et al. Potentially avoidable emergency department attendance: interview study of patients’ reasons for attendance. Emergency Medicine Journal 2012;29:e3.

[24] Kangovi, S., Barg, F. K., Carter, T., Long, J. A., Shannon, R., & Grande, D. (2013). Understanding why patients of low socioeconomic status prefer hospitals over ambulatory care. Health affairs, 32(7), 1196–1203.

[25] Edwards P, Green J, Lachowycz K, et al. Serious injuries in children: variation by area deprivation and settlement type. Archives of Disease in Childhood 2008;93:485–489.

[26] Groom L, Kendrick D, Coupland C, et al. Inequalities in hospital admission rates for unintentional poisoning in young children. Injury Prevention. 2006;12(3):166–70.

[27] Nussbaum, C., Massou, E., Fisher, R., Morciano, M., Harmer, R., & Ford, J. B. (2021). Inequalities in the distribution of the general practice workforce in England: a practice-level longitudinal analysis. BJGP Open, 5 (5), BJGPO. 2021.0066.

[28] Parisi, R., Lau, Y. S., Bower, P., Checkland, K., Rubery, J., Sutton, M.,… & Kontopantelis, E. (2023). Predictors and population health outcomes of persistent high GP turnover in English general practices: a retrospective observational study. BMJ Quality & Safety, 32(7), 394–403.

