## Supplementary table for "Inequalities in Accident and Emergency department attendance by socio-economic characteristics: population based study"

**Supplementary table 1. Comorbidities data for full sample population.**

|  | **Without comorbidities** | | **With comorbidities** | |
| --- | --- | --- | --- | --- |
| **Variable** | **Number** | **Percent** | **Number** | **Percent** |
| COVID-19 | 51,549,160.0 | 99.6% | 227,800 | 0.4% |
| Cancer | 49,282,925.0 | 95.2% | 2,494,030 | 4.8% |
| Diabetes | 50,121,765.0 | 96.8% | 1,655,190 | 3.2% |
| Dementia | 51,525,865.0 | 99.5% | 251,095 | 0.5% |
| Serious mental illness | 50,237,635.0 | 97.0% | 1,539,325 | 3.0% |
| Learning disability/autism | 51,670,190.0 | 99.8% | 106,765 | 0.2% |
| Neurological/Motor-nuerons Disease/Parkinsons/Multiple Sclerosis | 51,623,175.0 | 99.7% | 153,785 | 0.3% |
| Alzheimers | 51,674,090.0 | 99.8% | 102,870 | 0.2% |
| Epilepsy | 51,475,135.0 | 99.4% | 301,825 | 0.6% |
| Hypertension | 47,610,095.0 | 92.0% | 4,166,865 | 8.0% |
| Angina | 51,182,610.0 | 98.9% | 594,345 | 1.1% |
| Myocardial infarction | 51,532,780.0 | 99.5% | 244,180 | 0.5% |
| Ischaemic heart disease | 50,527,015.0 | 97.6% | 1,249,940 | 2.4% |
| Atrial fibrillation | 50,702,310.0 | 97.9% | 1,074,650 | 2.1% |
| Heart failure | 51,271,545.0 | 99.0% | 505,415 | 1.0% |
| Stroke | 51,557,215.0 | 99.6% | 219,745 | 0.4% |
| Other respiratory infection | 50,743,120.0 | 98.0% | 1,033,835 | 2.0% |
| Influenza/Pneumonia | 50,988,660.0 | 98.5% | 788,300 | 1.5% |
| COPD_respitory_failure | 50,977,965.0 | 98.5% | 798,990 | 1.5% |
| Asthma | 49,863,680.0 | 96.3% | 1,913,275 | 3.7% |
| Bronchiectasis | 51,643,160.0 | 99.7% | 133,795 | 0.3% |
| Inflammatory bowel disease | 50,942,620.0 | 98.4% | 834,335 | 1.6% |
| Liver disease | 51,350,875.0 | 99.2% | 426,085 | 0.8% |
| Rheumatoid arthritis | 51,473,020.0 | 99.4% | 303,935 | 0.6% |
| Oesteoarthritis | 49,983,095.0 | 96.5% | 1,793,860 | 3.5% |
| Oesteoporosis | 51,360,140.0 | 99.2% | 416,815 | 0.8% |
| Kidney disease | 50,202,805.0 | 97.0% | 1,574,155 | 3.0% |
| Respiratory Cancer | 51,715,325.0 | 99.9% | 61,635 | 0.1% |
| Accident and Emergency Attendance | 40,278,440.0 | 77.8% | 11,498,520 | 22.2% |
| Accident and Emergency Attendance - Immediate acuity | 51,563,530.0 | 99.6% | 213,430 | 0.4% |
| Accident and Emergency Attendance - Very Urgent acuity | 50,621,980.0 | 97.8% | 1,154,980 | 2.2% |
| Accident and Emergency Attendance - Urgent acuity | 47,794,725.0 | 92.3% | 3,982,230 | 7.7% |
| Accident and Emergency Attendance - Standard acuity | 45,930,280.0 | 88.7% | 5,846,680 | 11.3% |
| Accident and Emergency Attendance - Low acuity | 50,709,055.0 | 97.9% | 1,067,900 | 2.1% |

**Supplementary table 2. Odds ratios by exposure and age for unadjusted and health-adjusted models. Reference categories are IMD decile 10, Higher managerial, administrative and professional occupations and Highest qualification level 4: Degree and above for IMD, NS-SEC and qualifications respectively.**

| **Level** | **Model adjustment** | **Age group** | **Odds ratio** | **Odds ratio lower 95%** | **Odds ratio upper 95%** | **P-value** |
| --- | --- | --- | --- | --- | --- | --- |
| IMD Decile 1 | Not health adjusted | 0 to 5 years old | 1.56 | 1.54 | 1.57 | 0 |
| IMD Decile 1 | Health adjusted | 0 to 5 years old | 1.5 | 1.48 | 1.51 | 0 |
| IMD Decile 1 | Not health adjusted | 6 to 15 years old | 1.46 | 1.45 | 1.47 | 0 |
| IMD Decile 1 | Health adjusted | 6 to 15 years old | 1.4 | 1.38 | 1.41 | 0 |
| IMD Decile 1 | Not health adjusted | 16 to 29 years old | 1.57 | 1.56 | 1.58 | 0 |
| IMD Decile 1 | Health adjusted | 16 to 29 years old | 1.43 | 1.42 | 1.44 | 0 |
| IMD Decile 1 | Not health adjusted | 30 to 49 years old | 1.86 | 1.85 | 1.87 | 0 |
| IMD Decile 1 | Health adjusted | 30 to 49 years old | 1.54 | 1.53 | 1.55 | 0 |
| IMD Decile 1 | Not health adjusted | 50 to 64 years old | 1.87 | 1.86 | 1.88 | 0 |
| IMD Decile 1 | Health adjusted | 50 to 64 years old | 1.39 | 1.38 | 1.4 | 0 |
| IMD Decile 1 | Not health adjusted | 65 to 79 years old | 1.73 | 1.71 | 1.74 | 0 |
| IMD Decile 1 | Health adjusted | 65 to 79 years old | 1.23 | 1.22 | 1.24 | 0 |
| IMD Decile 1 | Not health adjusted | 80 to 95 years old | 1.46 | 1.45 | 1.48 | 0 |
| IMD Decile 1 | Health adjusted | 80 to 95 years old | 1.19 | 1.18 | 1.21 | 0 |
| IMD Decile 2 | Not health adjusted | 0 to 5 years old | 1.45 | 1.43 | 1.46 | 0 |
| IMD Decile 2 | Health adjusted | 0 to 5 years old | 1.4 | 1.39 | 1.42 | 0 |
| IMD Decile 2 | Not health adjusted | 6 to 15 years old | 1.36 | 1.35 | 1.37 | 0 |
| IMD Decile 2 | Health adjusted | 6 to 15 years old | 1.31 | 1.3 | 1.32 | 0 |
| IMD Decile 2 | Not health adjusted | 16 to 29 years old | 1.42 | 1.41 | 1.43 | 0 |
| IMD Decile 2 | Health adjusted | 16 to 29 years old | 1.34 | 1.33 | 1.35 | 0 |
| IMD Decile 2 | Not health adjusted | 30 to 49 years old | 1.66 | 1.65 | 1.67 | 0 |
| IMD Decile 2 | Health adjusted | 30 to 49 years old | 1.44 | 1.43 | 1.45 | 0 |
| IMD Decile 2 | Not health adjusted | 50 to 64 years old | 1.7 | 1.69 | 1.71 | 0 |
| IMD Decile 2 | Health adjusted | 50 to 64 years old | 1.34 | 1.33 | 1.35 | 0 |
| IMD Decile 2 | Not health adjusted | 65 to 79 years old | 1.57 | 1.56 | 1.58 | 0 |
| IMD Decile 2 | Health adjusted | 65 to 79 years old | 1.2 | 1.19 | 1.21 | 0 |
| IMD Decile 2 | Not health adjusted | 80 to 95 years old | 1.38 | 1.36 | 1.39 | 0 |
| IMD Decile 2 | Health adjusted | 80 to 95 years old | 1.17 | 1.15 | 1.18 | 0 |
| IMD Decile 3 | Not health adjusted | 0 to 5 years old | 1.36 | 1.35 | 1.38 | 0 |
| IMD Decile 3 | Health adjusted | 0 to 5 years old | 1.33 | 1.32 | 1.35 | 0 |
| IMD Decile 3 | Not health adjusted | 6 to 15 years old | 1.29 | 1.28 | 1.3 | 0 |
| IMD Decile 3 | Health adjusted | 6 to 15 years old | 1.25 | 1.24 | 1.26 | 0 |
| IMD Decile 3 | Not health adjusted | 16 to 29 years old | 1.33 | 1.32 | 1.34 | 0 |
| IMD Decile 3 | Health adjusted | 16 to 29 years old | 1.27 | 1.26 | 1.28 | 0 |
| IMD Decile 3 | Not health adjusted | 30 to 49 years old | 1.51 | 1.5 | 1.52 | 0 |
| IMD Decile 3 | Health adjusted | 30 to 49 years old | 1.35 | 1.34 | 1.36 | 0 |
| IMD Decile 3 | Not health adjusted | 50 to 64 years old | 1.56 | 1.54 | 1.57 | 0 |
| IMD Decile 3 | Health adjusted | 50 to 64 years old | 1.28 | 1.27 | 1.29 | 0 |
| IMD Decile 3 | Not health adjusted | 65 to 79 years old | 1.44 | 1.43 | 1.46 | 0 |
| IMD Decile 3 | Health adjusted | 65 to 79 years old | 1.16 | 1.15 | 1.17 | 0 |
| IMD Decile 3 | Not health adjusted | 80 to 95 years old | 1.31 | 1.29 | 1.32 | 0 |
| IMD Decile 3 | Health adjusted | 80 to 95 years old | 1.14 | 1.12 | 1.15 | 0 |
| IMD Decile 4 | Not health adjusted | 0 to 5 years old | 1.29 | 1.28 | 1.31 | 0 |
| IMD Decile 4 | Health adjusted | 0 to 5 years old | 1.27 | 1.25 | 1.28 | 0 |
| IMD Decile 4 | Not health adjusted | 6 to 15 years old | 1.24 | 1.23 | 1.25 | 0 |
| IMD Decile 4 | Health adjusted | 6 to 15 years old | 1.2 | 1.19 | 1.21 | 0 |
| IMD Decile 4 | Not health adjusted | 16 to 29 years old | 1.25 | 1.24 | 1.26 | 0 |
| IMD Decile 4 | Health adjusted | 16 to 29 years old | 1.21 | 1.2 | 1.22 | 0 |
| IMD Decile 4 | Not health adjusted | 30 to 49 years old | 1.41 | 1.41 | 1.42 | 0 |
| IMD Decile 4 | Health adjusted | 30 to 49 years old | 1.29 | 1.28 | 1.3 | 0 |
| IMD Decile 4 | Not health adjusted | 50 to 64 years old | 1.43 | 1.42 | 1.44 | 0 |
| IMD Decile 4 | Health adjusted | 50 to 64 years old | 1.23 | 1.22 | 1.24 | 0 |
| IMD Decile 4 | Not health adjusted | 65 to 79 years old | 1.32 | 1.31 | 1.33 | 0 |
| IMD Decile 4 | Health adjusted | 65 to 79 years old | 1.12 | 1.11 | 1.13 | 0 |
| IMD Decile 4 | Not health adjusted | 80 to 95 years old | 1.2 | 1.19 | 1.22 | 0 |
| IMD Decile 4 | Health adjusted | 80 to 95 years old | 1.08 | 1.07 | 1.09 | 0 |
| IMD Decile 5 | Not health adjusted | 0 to 5 years old | 1.21 | 1.2 | 1.23 | 0 |
| IMD Decile 5 | Health adjusted | 0 to 5 years old | 1.2 | 1.18 | 1.21 | 0 |
| IMD Decile 5 | Not health adjusted | 6 to 15 years old | 1.16 | 1.15 | 1.17 | 0 |
| IMD Decile 5 | Health adjusted | 6 to 15 years old | 1.14 | 1.13 | 1.15 | 0 |
| IMD Decile 5 | Not health adjusted | 16 to 29 years old | 1.2 | 1.19 | 1.21 | 0 |
| IMD Decile 5 | Health adjusted | 16 to 29 years old | 1.16 | 1.15 | 1.17 | 0 |
| IMD Decile 5 | Not health adjusted | 30 to 49 years old | 1.32 | 1.31 | 1.33 | 0 |
| IMD Decile 5 | Health adjusted | 30 to 49 years old | 1.22 | 1.21 | 1.23 | 0 |
| IMD Decile 5 | Not health adjusted | 50 to 64 years old | 1.32 | 1.31 | 1.33 | 0 |
| IMD Decile 5 | Health adjusted | 50 to 64 years old | 1.17 | 1.16 | 1.18 | 0 |
| IMD Decile 5 | Not health adjusted | 65 to 79 years old | 1.22 | 1.21 | 1.23 | 0 |
| IMD Decile 5 | Health adjusted | 65 to 79 years old | 1.08 | 1.07 | 1.09 | 0 |
| IMD Decile 5 | Not health adjusted | 80 to 95 years old | 1.14 | 1.13 | 1.16 | 0 |
| IMD Decile 5 | Health adjusted | 80 to 95 years old | 1.05 | 1.04 | 1.06 | 0 |
| IMD Decile 6 | Not health adjusted | 0 to 5 years old | 1.16 | 1.15 | 1.18 | 0 |
| IMD Decile 6 | Health adjusted | 0 to 5 years old | 1.15 | 1.14 | 1.16 | 0 |
| IMD Decile 6 | Not health adjusted | 6 to 15 years old | 1.13 | 1.12 | 1.14 | 0 |
| IMD Decile 6 | Health adjusted | 6 to 15 years old | 1.11 | 1.1 | 1.12 | 0 |
| IMD Decile 6 | Not health adjusted | 16 to 29 years old | 1.16 | 1.15 | 1.17 | 0 |
| IMD Decile 6 | Health adjusted | 16 to 29 years old | 1.14 | 1.13 | 1.15 | 0 |
| IMD Decile 6 | Not health adjusted | 30 to 49 years old | 1.25 | 1.24 | 1.26 | 0 |
| IMD Decile 6 | Health adjusted | 30 to 49 years old | 1.18 | 1.17 | 1.19 | 0 |
| IMD Decile 6 | Not health adjusted | 50 to 64 years old | 1.24 | 1.23 | 1.25 | 0 |
| IMD Decile 6 | Health adjusted | 50 to 64 years old | 1.13 | 1.12 | 1.14 | 0 |
| IMD Decile 6 | Not health adjusted | 65 to 79 years old | 1.17 | 1.16 | 1.18 | 0 |
| IMD Decile 6 | Health adjusted | 65 to 79 years old | 1.06 | 1.06 | 1.07 | 0 |
| IMD Decile 6 | Not health adjusted | 80 to 95 years old | 1.11 | 1.1 | 1.12 | 0 |
| IMD Decile 6 | Health adjusted | 80 to 95 years old | 1.04 | 1.03 | 1.05 | 0 |
| IMD Decile 7 | Not health adjusted | 0 to 5 years old | 1.13 | 1.12 | 1.15 | 0 |
| IMD Decile 7 | Health adjusted | 0 to 5 years old | 1.12 | 1.11 | 1.13 | 0 |
| IMD Decile 7 | Not health adjusted | 6 to 15 years old | 1.1 | 1.09 | 1.11 | 0 |
| IMD Decile 7 | Health adjusted | 6 to 15 years old | 1.08 | 1.08 | 1.09 | 0 |
| IMD Decile 7 | Not health adjusted | 16 to 29 years old | 1.12 | 1.11 | 1.13 | 0 |
| IMD Decile 7 | Health adjusted | 16 to 29 years old | 1.11 | 1.1 | 1.11 | 0 |
| IMD Decile 7 | Not health adjusted | 30 to 49 years old | 1.2 | 1.19 | 1.21 | 0 |
| IMD Decile 7 | Health adjusted | 30 to 49 years old | 1.15 | 1.14 | 1.15 | 0 |
| IMD Decile 7 | Not health adjusted | 50 to 64 years old | 1.19 | 1.18 | 1.19 | 0 |
| IMD Decile 7 | Health adjusted | 50 to 64 years old | 1.11 | 1.1 | 1.12 | 0 |
| IMD Decile 7 | Not health adjusted | 65 to 79 years old | 1.14 | 1.13 | 1.15 | 0 |
| IMD Decile 7 | Health adjusted | 65 to 79 years old | 1.06 | 1.05 | 1.07 | 0 |
| IMD Decile 7 | Not health adjusted | 80 to 95 years old | 1.1 | 1.09 | 1.11 | 0 |
| IMD Decile 7 | Health adjusted | 80 to 95 years old | 1.04 | 1.03 | 1.05 | 0 |
| IMD Decile 8 | Not health adjusted | 0 to 5 years old | 1.11 | 1.1 | 1.13 | 0 |
| IMD Decile 8 | Health adjusted | 0 to 5 years old | 1.11 | 1.09 | 1.12 | 0 |
| IMD Decile 8 | Not health adjusted | 6 to 15 years old | 1.09 | 1.08 | 1.1 | 0 |
| IMD Decile 8 | Health adjusted | 6 to 15 years old | 1.08 | 1.07 | 1.09 | 0 |
| IMD Decile 8 | Not health adjusted | 16 to 29 years old | 1.1 | 1.09 | 1.11 | 0 |
| IMD Decile 8 | Health adjusted | 16 to 29 years old | 1.09 | 1.08 | 1.1 | 0 |
| IMD Decile 8 | Not health adjusted | 30 to 49 years old | 1.16 | 1.15 | 1.17 | 0 |
| IMD Decile 8 | Health adjusted | 30 to 49 years old | 1.12 | 1.11 | 1.13 | 0 |
| IMD Decile 8 | Not health adjusted | 50 to 64 years old | 1.15 | 1.14 | 1.16 | 0 |
| IMD Decile 8 | Health adjusted | 50 to 64 years old | 1.1 | 1.09 | 1.1 | 0 |
| IMD Decile 8 | Not health adjusted | 65 to 79 years old | 1.12 | 1.11 | 1.12 | 0 |
| IMD Decile 8 | Health adjusted | 65 to 79 years old | 1.05 | 1.04 | 1.06 | 0 |
| IMD Decile 8 | Not health adjusted | 80 to 95 years old | 1.08 | 1.07 | 1.1 | 0 |
| IMD Decile 8 | Health adjusted | 80 to 95 years old | 1.04 | 1.03 | 1.05 | 0 |
| IMD Decile 9 | Not health adjusted | 0 to 5 years old | 1.07 | 1.06 | 1.08 | 0 |
| IMD Decile 9 | Health adjusted | 0 to 5 years old | 1.06 | 1.05 | 1.07 | 0 |
| IMD Decile 9 | Not health adjusted | 6 to 15 years old | 1.05 | 1.05 | 1.06 | 0 |
| IMD Decile 9 | Health adjusted | 6 to 15 years old | 1.05 | 1.04 | 1.06 | 0 |
| IMD Decile 9 | Not health adjusted | 16 to 29 years old | 1.07 | 1.06 | 1.08 | 0 |
| IMD Decile 9 | Health adjusted | 16 to 29 years old | 1.07 | 1.06 | 1.08 | 0 |
| IMD Decile 9 | Not health adjusted | 30 to 49 years old | 1.1 | 1.09 | 1.11 | 0 |
| IMD Decile 9 | Health adjusted | 30 to 49 years old | 1.08 | 1.07 | 1.08 | 0 |
| IMD Decile 9 | Not health adjusted | 50 to 64 years old | 1.09 | 1.09 | 1.1 | 0 |
| IMD Decile 9 | Health adjusted | 50 to 64 years old | 1.06 | 1.05 | 1.07 | 0 |
| IMD Decile 9 | Not health adjusted | 65 to 79 years old | 1.07 | 1.06 | 1.07 | 0 |
| IMD Decile 9 | Health adjusted | 65 to 79 years old | 1.03 | 1.02 | 1.04 | 0 |
| IMD Decile 9 | Not health adjusted | 80 to 95 years old | 1.04 | 1.03 | 1.05 | 0 |
| IMD Decile 9 | Health adjusted | 80 to 95 years old | 1.01 | 1 | 1.02 | 0.04 |
| NS-SEC 2 Lower managerial, administrative and professional occupations | Not health adjusted | 25 to 49 years old | 1.22 | 1.22 | 1.23 | 0 |
| NS-SEC 2 Lower managerial, administrative and professional occupations | Health adjusted | 25 to 49 years old | 1.18 | 1.17 | 1.18 | 0 |
| NS-SEC 2 Lower managerial, administrative and professional occupations | Not health adjusted | 50 to 64 years old | 1.21 | 1.2 | 1.21 | 0 |
| NS-SEC 2 Lower managerial, administrative and professional occupations | Health adjusted | 50 to 64 years old | 1.14 | 1.13 | 1.15 | 0 |
| NS-SEC 2 Lower managerial, administrative and professional occupations | Not health adjusted | 65 to 79 years old | 1.12 | 1.11 | 1.13 | 0 |
| NS-SEC 2 Lower managerial, administrative and professional occupations | Health adjusted | 65 to 79 years old | 1.05 | 1.04 | 1.06 | 0 |
| NS-SEC 2 Lower managerial, administrative and professional occupations | Not health adjusted | 80 to 95 years old | 1.09 | 1.08 | 1.1 | 0 |
| NS-SEC 2 Lower managerial, administrative and professional occupations | Health adjusted | 80 to 95 years old | 1.04 | 1.03 | 1.05 | 0 |
| NS-SEC 3 Intermediate occupations | Not health adjusted | 25 to 49 years old | 1.39 | 1.39 | 1.4 | 0 |
| NS-SEC 3 Intermediate occupations | Health adjusted | 25 to 49 years old | 1.27 | 1.27 | 1.28 | 0 |
| NS-SEC 3 Intermediate occupations | Not health adjusted | 50 to 64 years old | 1.26 | 1.25 | 1.27 | 0 |
| NS-SEC 3 Intermediate occupations | Health adjusted | 50 to 64 years old | 1.15 | 1.14 | 1.15 | 0 |
| NS-SEC 3 Intermediate occupations | Not health adjusted | 65 to 79 years old | 1.14 | 1.13 | 1.14 | 0 |
| NS-SEC 3 Intermediate occupations | Health adjusted | 65 to 79 years old | 1.05 | 1.04 | 1.06 | 0 |
| NS-SEC 3 Intermediate occupations | Not health adjusted | 80 to 95 years old | 1.11 | 1.1 | 1.12 | 0 |
| NS-SEC 3 Intermediate occupations | Health adjusted | 80 to 95 years old | 1.05 | 1.04 | 1.07 | 0 |
| NS-SEC 4 Small employers and own account workers | Not health adjusted | 25 to 49 years old | 1.52 | 1.51 | 1.52 | 0 |
| NS-SEC 4 Small employers and own account workers | Health adjusted | 25 to 49 years old | 1.45 | 1.44 | 1.46 | 0 |
| NS-SEC 4 Small employers and own account workers | Not health adjusted | 50 to 64 years old | 1.37 | 1.36 | 1.37 | 0 |
| NS-SEC 4 Small employers and own account workers | Health adjusted | 50 to 64 years old | 1.26 | 1.25 | 1.27 | 0 |
| NS-SEC 4 Small employers and own account workers | Not health adjusted | 65 to 79 years old | 1.26 | 1.25 | 1.27 | 0 |
| NS-SEC 4 Small employers and own account workers | Health adjusted | 65 to 79 years old | 1.14 | 1.13 | 1.14 | 0 |
| NS-SEC 4 Small employers and own account workers | Not health adjusted | 80 to 95 years old | 1.17 | 1.16 | 1.18 | 0 |
| NS-SEC 4 Small employers and own account workers | Health adjusted | 80 to 95 years old | 1.08 | 1.06 | 1.09 | 0 |
| NS-SEC 5 Lower supervisory and technical occupations | Not health adjusted | 25 to 49 years old | 1.6 | 1.59 | 1.61 | 0 |
| NS-SEC 5 Lower supervisory and technical occupations | Health adjusted | 25 to 49 years old | 1.48 | 1.47 | 1.49 | 0 |
| NS-SEC 5 Lower supervisory and technical occupations | Not health adjusted | 50 to 64 years old | 1.5 | 1.49 | 1.51 | 0 |
| NS-SEC 5 Lower supervisory and technical occupations | Health adjusted | 50 to 64 years old | 1.29 | 1.28 | 1.3 | 0 |
| NS-SEC 5 Lower supervisory and technical occupations | Not health adjusted | 65 to 79 years old | 1.3 | 1.29 | 1.32 | 0 |
| NS-SEC 5 Lower supervisory and technical occupations | Health adjusted | 65 to 79 years old | 1.08 | 1.06 | 1.09 | 0 |
| NS-SEC 5 Lower supervisory and technical occupations | Not health adjusted | 80 to 95 years old | 1.23 | 1.22 | 1.25 | 0 |
| NS-SEC 5 Lower supervisory and technical occupations | Health adjusted | 80 to 95 years old | 1.08 | 1.07 | 1.1 | 0 |
| NS-SEC 6 Semi-routine occupations | Not health adjusted | 25 to 49 years old | 1.69 | 1.68 | 1.7 | 0 |
| NS-SEC 6 Semi-routine occupations | Health adjusted | 25 to 49 years old | 1.44 | 1.43 | 1.45 | 0 |
| NS-SEC 6 Semi-routine occupations | Not health adjusted | 50 to 64 years old | 1.57 | 1.56 | 1.58 | 0 |
| NS-SEC 6 Semi-routine occupations | Health adjusted | 50 to 64 years old | 1.3 | 1.29 | 1.31 | 0 |
| NS-SEC 6 Semi-routine occupations | Not health adjusted | 65 to 79 years old | 1.31 | 1.3 | 1.32 | 0 |
| NS-SEC 6 Semi-routine occupations | Health adjusted | 65 to 79 years old | 1.1 | 1.09 | 1.11 | 0 |
| NS-SEC 6 Semi-routine occupations | Not health adjusted | 80 to 95 years old | 1.21 | 1.2 | 1.23 | 0 |
| NS-SEC 6 Semi-routine occupations | Health adjusted | 80 to 95 years old | 1.08 | 1.07 | 1.09 | 0 |
| NS-SEC 7 Routine occupations | Not health adjusted | 25 to 49 years old | 1.7 | 1.69 | 1.71 | 0 |
| NS-SEC 7 Routine occupations | Health adjusted | 25 to 49 years old | 1.5 | 1.49 | 1.5 | 0 |
| NS-SEC 7 Routine occupations | Not health adjusted | 50 to 64 years old | 1.62 | 1.61 | 1.63 | 0 |
| NS-SEC 7 Routine occupations | Health adjusted | 50 to 64 years old | 1.33 | 1.32 | 1.34 | 0 |
| NS-SEC 7 Routine occupations | Not health adjusted | 65 to 79 years old | 1.42 | 1.41 | 1.43 | 0 |
| NS-SEC 7 Routine occupations | Health adjusted | 65 to 79 years old | 1.13 | 1.12 | 1.14 | 0 |
| NS-SEC 7 Routine occupations | Not health adjusted | 80 to 95 years old | 1.3 | 1.29 | 1.32 | 0 |
| NS-SEC 7 Routine occupations | Health adjusted | 80 to 95 years old | 1.11 | 1.1 | 1.12 | 0 |
| NS-SEC 8 Never worked and long-term unemployed | Not health adjusted | 25 to 49 years old | 1.96 | 1.95 | 1.97 | 0 |
| NS-SEC 8 Never worked and long-term unemployed | Health adjusted | 25 to 49 years old | 1.41 | 1.4 | 1.42 | 0 |
| NS-SEC 8 Never worked and long-term unemployed | Not health adjusted | 50 to 64 years old | 2.09 | 2.07 | 2.1 | 0 |
| NS-SEC 8 Never worked and long-term unemployed | Health adjusted | 50 to 64 years old | 1.32 | 1.31 | 1.33 | 0 |
| NS-SEC 8 Never worked and long-term unemployed | Not health adjusted | 65 to 79 years old | 1.59 | 1.58 | 1.6 | 0 |
| NS-SEC 8 Never worked and long-term unemployed | Health adjusted | 65 to 79 years old | 1.21 | 1.2 | 1.22 | 0 |
| NS-SEC 8 Never worked and long-term unemployed | Not health adjusted | 80 to 95 years old | 1.29 | 1.28 | 1.31 | 0 |
| NS-SEC 8 Never worked and long-term unemployed | Health adjusted | 80 to 95 years old | 1.12 | 1.11 | 1.13 | 0 |
| Highest qualification level 1: 1-4 GCSEs and equivalent | Not health adjusted | 25 to 49 years old | 1.49 | 1.48 | 1.5 | 0 |
| Highest qualification level 1: 1-4 GCSEs and equivalent | Health adjusted | 25 to 49 years old | 1.3 | 1.29 | 1.3 | 0 |
| Highest qualification level 1: 1-4 GCSEs and equivalent | Not health adjusted | 50 to 64 years old | 1.21 | 1.2 | 1.21 | 0 |
| Highest qualification level 1: 1-4 GCSEs and equivalent | Health adjusted | 50 to 64 years old | 1.08 | 1.07 | 1.08 | 0 |
| Highest qualification level 1: 1-4 GCSEs and equivalent | Not health adjusted | 65 to 79 years old | 1.14 | 1.13 | 1.14 | 0 |
| Highest qualification level 1: 1-4 GCSEs and equivalent | Health adjusted | 65 to 79 years old | 1.03 | 1.02 | 1.03 | 0 |
| Highest qualification level 1: 1-4 GCSEs and equivalent | Not health adjusted | 80 to 95 years old | 1.15 | 1.14 | 1.16 | 0 |
| Highest qualification level 1: 1-4 GCSEs and equivalent | Health adjusted | 80 to 95 years old | 1.05 | 1.04 | 1.07 | 0 |
| Highest qualification level 2: 5+ GCSEs and equivalent | Not health adjusted | 25 to 49 years old | 1.4 | 1.39 | 1.4 | 0 |
| Highest qualification level 2: 5+ GCSEs and equivalent | Health adjusted | 25 to 49 years old | 1.26 | 1.26 | 1.27 | 0 |
| Highest qualification level 2: 5+ GCSEs and equivalent | Not health adjusted | 50 to 64 years old | 1.16 | 1.15 | 1.16 | 0 |
| Highest qualification level 2: 5+ GCSEs and equivalent | Health adjusted | 50 to 64 years old | 1.07 | 1.06 | 1.07 | 0 |
| Highest qualification level 2: 5+ GCSEs and equivalent | Not health adjusted | 65 to 79 years old | 1.06 | 1.05 | 1.07 | 0 |
| Highest qualification level 2: 5+ GCSEs and equivalent | Health adjusted | 65 to 79 years old | 1 | 1 | 1.01 | 0.34 |
| Highest qualification level 2: 5+ GCSEs and equivalent | Not health adjusted | 80 to 95 years old | 1.02 | 1.01 | 1.04 | 0 |
| Highest qualification level 2: 5+ GCSEs and equivalent | Health adjusted | 80 to 95 years old | 1.01 | 1 | 1.02 | 0.2 |
| Highest qualification level 3: A levels, AS levels and equivalent | Not health adjusted | 25 to 49 years old | 1.26 | 1.26 | 1.27 | 0 |
| Highest qualification level 3: A levels, AS levels and equivalent | Health adjusted | 25 to 49 years old | 1.19 | 1.18 | 1.19 | 0 |
| Highest qualification level 3: A levels, AS levels and equivalent | Not health adjusted | 50 to 64 years old | 1.14 | 1.14 | 1.15 | 0 |
| Highest qualification level 3: A levels, AS levels and equivalent | Health adjusted | 50 to 64 years old | 1.08 | 1.08 | 1.09 | 0 |
| Highest qualification level 3: A levels, AS levels and equivalent | Not health adjusted | 65 to 79 years old | 1.09 | 1.09 | 1.1 | 0 |
| Highest qualification level 3: A levels, AS levels and equivalent | Health adjusted | 65 to 79 years old | 1.03 | 1.02 | 1.04 | 0 |
| Highest qualification level 3: A levels, AS levels and equivalent | Not health adjusted | 80 to 95 years old | 1.06 | 1.04 | 1.07 | 0 |
| Highest qualification level 3: A levels, AS levels and equivalent | Health adjusted | 80 to 95 years old | 1.02 | 1 | 1.03 | 0.02 |
| Highest qualification: Apprenticeship | Not health adjusted | 25 to 49 years old | 1.47 | 1.46 | 1.48 | 0 |
| Highest qualification: Apprenticeship | Health adjusted | 25 to 49 years old | 1.37 | 1.36 | 1.37 | 0 |
| Highest qualification: Apprenticeship | Not health adjusted | 50 to 64 years old | 1.27 | 1.26 | 1.28 | 0 |
| Highest qualification: Apprenticeship | Health adjusted | 50 to 64 years old | 1.15 | 1.14 | 1.16 | 0 |
| Highest qualification: Apprenticeship | Not health adjusted | 65 to 79 years old | 1.18 | 1.17 | 1.19 | 0 |
| Highest qualification: Apprenticeship | Health adjusted | 65 to 79 years old | 1.05 | 1.04 | 1.05 | 0 |
| Highest qualification: Apprenticeship | Not health adjusted | 80 to 95 years old | 1.14 | 1.13 | 1.15 | 0 |
| Highest qualification: Apprenticeship | Health adjusted | 80 to 95 years old | 1.04 | 1.03 | 1.05 | 0 |
| Highest qualification: no qualification | Not health adjusted | 25 to 49 years old | 1.62 | 1.62 | 1.63 | 0 |
| Highest qualification: no qualification | Health adjusted | 25 to 49 years old | 1.37 | 1.36 | 1.38 | 0 |
| Highest qualification: no qualification | Not health adjusted | 50 to 64 years old | 1.57 | 1.56 | 1.58 | 0 |
| Highest qualification: no qualification | Health adjusted | 50 to 64 years old | 1.21 | 1.21 | 1.22 | 0 |
| Highest qualification: no qualification | Not health adjusted | 65 to 79 years old | 1.36 | 1.35 | 1.36 | 0 |
| Highest qualification: no qualification | Health adjusted | 65 to 79 years old | 1.09 | 1.09 | 1.1 | 0 |
| Highest qualification: no qualification | Not health adjusted | 80 to 95 years old | 1.23 | 1.22 | 1.24 | 0 |
| Highest qualification: no qualification | Health adjusted | 80 to 95 years old | 1.08 | 1.07 | 1.09 | 0 |

**Supplementary table 2. Odds ratios by exposure and acuity for unadjusted and health-adjusted models. Reference categories are IMD decile 10, Higher managerial, administrative, and professional occupations and Highest qualification level 4: Degree and above for IMD, NS-SEC and qualifications respectively.**

| **Acuity** | **Level** | **Model adjustment** | **Odds ratio** | **Odds ratio lower 95%** | **Odds ratio upper 95%** | **P-value** |
| --- | --- | --- | --- | --- | --- | --- |
| All | IMD Decile 1 | Not health adjusted | 1.69 | 1.68 | 1.69 | 0 |
| All | IMD Decile 1 | Health adjusted | 1.41 | 1.4 | 1.41 | 0 |
| All | IMD Decile 2 | Not health adjusted | 1.55 | 1.54 | 1.55 | 0 |
| All | IMD Decile 2 | Health adjusted | 1.34 | 1.33 | 1.34 | 0 |
| All | IMD Decile 3 | Not health adjusted | 1.44 | 1.44 | 1.44 | 0 |
| All | IMD Decile 3 | Health adjusted | 1.27 | 1.27 | 1.28 | 0 |
| All | IMD Decile 4 | Not health adjusted | 1.35 | 1.34 | 1.35 | 0 |
| All | IMD Decile 4 | Health adjusted | 1.22 | 1.21 | 1.22 | 0 |
| All | IMD Decile 5 | Not health adjusted | 1.26 | 1.25 | 1.26 | 0 |
| All | IMD Decile 5 | Health adjusted | 1.16 | 1.16 | 1.16 | 0 |
| All | IMD Decile 6 | Not health adjusted | 1.2 | 1.2 | 1.2 | 0 |
| All | IMD Decile 6 | Health adjusted | 1.13 | 1.12 | 1.13 | 0 |
| All | IMD Decile 7 | Not health adjusted | 1.16 | 1.15 | 1.16 | 0 |
| All | IMD Decile 7 | Health adjusted | 1.1 | 1.1 | 1.11 | 0 |
| All | IMD Decile 8 | Not health adjusted | 1.13 | 1.12 | 1.13 | 0 |
| All | IMD Decile 8 | Health adjusted | 1.09 | 1.09 | 1.09 | 0 |
| All | IMD Decile 9 | Not health adjusted | 1.08 | 1.08 | 1.08 | 0 |
| All | IMD Decile 9 | Health adjusted | 1.06 | 1.05 | 1.06 | 0 |
| All | NS-SEC 2 Lower managerial, administrative, and professional occupations | Not health adjusted | 1.19 | 1.19 | 1.19 | 0 |
| All | NS-SEC 2 Lower managerial, administrative, and professional occupations | Health adjusted | 1.14 | 1.14 | 1.14 | 0 |
| All | NS-SEC 3 Intermediate occupations | Not health adjusted | 1.28 | 1.28 | 1.29 | 0 |
| All | NS-SEC 3 Intermediate occupations | Health adjusted | 1.19 | 1.19 | 1.19 | 0 |
| All | NS-SEC 4 Small employers and own account workers | Not health adjusted | 1.39 | 1.39 | 1.4 | 0 |
| All | NS-SEC 4 Small employers and own account workers | Health adjusted | 1.31 | 1.31 | 1.32 | 0 |
| All | NS-SEC 5 Lower supervisory and technical occupations | Not health adjusted | 1.5 | 1.49 | 1.5 | 0 |
| All | NS-SEC 5 Lower supervisory and technical occupations | Health adjusted | 1.34 | 1.34 | 1.35 | 0 |
| All | NS-SEC 6 Semi-routine occupations | Not health adjusted | 1.52 | 1.52 | 1.53 | 0 |
| All | NS-SEC 6 Semi-routine occupations | Health adjusted | 1.31 | 1.31 | 1.31 | 0 |
| All | NS-SEC 7 Routine occupations | Not health adjusted | 1.58 | 1.57 | 1.58 | 0 |
| All | NS-SEC 7 Routine occupations | Health adjusted | 1.35 | 1.35 | 1.36 | 0 |
| All | NS-SEC 8 Never worked and long-term unemployed | Not health adjusted | 1.74 | 1.74 | 1.75 | 0 |
| All | NS-SEC 8 Never worked and long-term unemployed | Health adjusted | 1.3 | 1.3 | 1.31 | 0 |
| All | Highest qualification level 1: 1-4 GCSEs and equivalent | Not health adjusted | 1.28 | 1.28 | 1.29 | 0 |
| All | Highest qualification level 1: 1-4 GCSEs and equivalent | Health adjusted | 1.16 | 1.16 | 1.16 | 0 |
| All | Highest qualification level 2: 5+ GCSEs and equivalent | Not health adjusted | 1.19 | 1.18 | 1.19 | 0 |
| All | Highest qualification level 2: 5+ GCSEs and equivalent | Health adjusted | 1.12 | 1.12 | 1.12 | 0 |
| All | Highest qualification level 3: A levels, AS levels and equivalent | Not health adjusted | 1.18 | 1.17 | 1.18 | 0 |
| All | Highest qualification level 3: A levels, AS levels and equivalent | Health adjusted | 1.12 | 1.12 | 1.12 | 0 |
| All | Highest qualification: Apprenticeship | Not health adjusted | 1.31 | 1.3 | 1.31 | 0 |
| All | Highest qualification: Apprenticeship | Health adjusted | 1.2 | 1.2 | 1.21 | 0 |
| All | Highest qualification: no qualification | Not health adjusted | 1.43 | 1.43 | 1.43 | 0 |
| All | Highest qualification: no qualification | Health adjusted | 1.2 | 1.19 | 1.2 | 0 |
| Low acuity | IMD Decile 1 | Not health adjusted | 2.26 | 2.23 | 2.28 | 0 |
| Low acuity | IMD Decile 1 | Health adjusted | 2.02 | 2 | 2.04 | 0 |
| Low acuity | IMD Decile 2 | Not health adjusted | 2.06 | 2.04 | 2.08 | 0 |
| Low acuity | IMD Decile 2 | Health adjusted | 1.89 | 1.87 | 1.91 | 0 |
| Low acuity | IMD Decile 3 | Not health adjusted | 1.83 | 1.81 | 1.85 | 0 |
| Low acuity | IMD Decile 3 | Health adjusted | 1.7 | 1.69 | 1.72 | 0 |
| Low acuity | IMD Decile 4 | Not health adjusted | 2.03 | 2.01 | 2.05 | 0 |
| Low acuity | IMD Decile 4 | Health adjusted | 1.92 | 1.9 | 1.94 | 0 |
| Low acuity | IMD Decile 5 | Not health adjusted | 1.81 | 1.8 | 1.83 | 0 |
| Low acuity | IMD Decile 5 | Health adjusted | 1.73 | 1.72 | 1.75 | 0 |
| Low acuity | IMD Decile 6 | Not health adjusted | 1.54 | 1.52 | 1.55 | 0 |
| Low acuity | IMD Decile 6 | Health adjusted | 1.49 | 1.47 | 1.5 | 0 |
| Low acuity | IMD Decile 7 | Not health adjusted | 1.5 | 1.48 | 1.51 | 0 |
| Low acuity | IMD Decile 7 | Health adjusted | 1.46 | 1.44 | 1.47 | 0 |
| Low acuity | IMD Decile 8 | Not health adjusted | 1.37 | 1.35 | 1.38 | 0 |
| Low acuity | IMD Decile 8 | Health adjusted | 1.34 | 1.33 | 1.35 | 0 |
| Low acuity | IMD Decile 9 | Not health adjusted | 1.26 | 1.25 | 1.27 | 0 |
| Low acuity | IMD Decile 9 | Health adjusted | 1.25 | 1.23 | 1.26 | 0 |
| Low acuity | NS-SEC 2 Lower managerial, administrative and professional occupations | Not health adjusted | 1.24 | 1.23 | 1.25 | 0 |
| Low acuity | NS-SEC 2 Lower managerial, administrative and professional occupations | Health adjusted | 1.2 | 1.19 | 1.21 | 0 |
| Low acuity | NS-SEC 3 Intermediate occupations | Not health adjusted | 1.34 | 1.33 | 1.35 | 0 |
| Low acuity | NS-SEC 3 Intermediate occupations | Health adjusted | 1.26 | 1.25 | 1.28 | 0 |
| Low acuity | NS-SEC 4 Small employers and own account workers | Not health adjusted | 1.52 | 1.5 | 1.53 | 0 |
| Low acuity | NS-SEC 4 Small employers and own account workers | Health adjusted | 1.45 | 1.44 | 1.47 | 0 |
| Low acuity | NS-SEC 5 Lower supervisory and technical occupations | Not health adjusted | 1.59 | 1.57 | 1.61 | 0 |
| Low acuity | NS-SEC 5 Lower supervisory and technical occupations | Health adjusted | 1.48 | 1.46 | 1.49 | 0 |
| Low acuity | NS-SEC 6 Semi-routine occupations | Not health adjusted | 1.65 | 1.63 | 1.66 | 0 |
| Low acuity | NS-SEC 6 Semi-routine occupations | Health adjusted | 1.47 | 1.46 | 1.49 | 0 |
| Low acuity | NS-SEC 7 Routine occupations | Not health adjusted | 1.63 | 1.61 | 1.65 | 0 |
| Low acuity | NS-SEC 7 Routine occupations | Health adjusted | 1.47 | 1.45 | 1.48 | 0 |
| Low acuity | NS-SEC 8 Never worked and long-term unemployed | Not health adjusted | 1.75 | 1.74 | 1.77 | 0 |
| Low acuity | NS-SEC 8 Never worked and long-term unemployed | Health adjusted | 1.43 | 1.41 | 1.44 | 0 |
| Low acuity | Highest qualification level 1: 1-4 GCSEs and equivalent | Not health adjusted | 1.31 | 1.3 | 1.32 | 0 |
| Low acuity | Highest qualification level 1: 1-4 GCSEs and equivalent | Health adjusted | 1.2 | 1.19 | 1.21 | 0 |
| Low acuity | Highest qualification level 2: 5+ GCSEs and equivalent | Not health adjusted | 1.23 | 1.22 | 1.24 | 0 |
| Low acuity | Highest qualification level 2: 5+ GCSEs and equivalent | Health adjusted | 1.16 | 1.15 | 1.17 | 0 |
| Low acuity | Highest qualification level 3: A levels, AS levels and equivalent | Not health adjusted | 1.2 | 1.2 | 1.21 | 0 |
| Low acuity | Highest qualification level 3: A levels, AS levels and equivalent | Health adjusted | 1.16 | 1.15 | 1.16 | 0 |
| Low acuity | Highest qualification: Apprenticeship | Not health adjusted | 1.33 | 1.32 | 1.35 | 0 |
| Low acuity | Highest qualification: Apprenticeship | Health adjusted | 1.25 | 1.24 | 1.26 | 0 |
| Low acuity | Highest qualification: no qualification | Not health adjusted | 1.34 | 1.33 | 1.34 | 0 |
| Low acuity | Highest qualification: no qualification | Health adjusted | 1.18 | 1.17 | 1.19 | 0 |
| Standard acuity | IMD Decile 1 | Not health adjusted | 1.66 | 1.65 | 1.66 | 0 |
| Standard acuity | IMD Decile 1 | Health adjusted | 1.46 | 1.46 | 1.47 | 0 |
| Standard acuity | IMD Decile 2 | Not health adjusted | 1.49 | 1.49 | 1.5 | 0 |
| Standard acuity | IMD Decile 2 | Health adjusted | 1.35 | 1.34 | 1.35 | 0 |
| Standard acuity | IMD Decile 3 | Not health adjusted | 1.4 | 1.39 | 1.41 | 0 |
| Standard acuity | IMD Decile 3 | Health adjusted | 1.29 | 1.28 | 1.29 | 0 |
| Standard acuity | IMD Decile 4 | Not health adjusted | 1.28 | 1.28 | 1.29 | 0 |
| Standard acuity | IMD Decile 4 | Health adjusted | 1.2 | 1.19 | 1.2 | 0 |
| Standard acuity | IMD Decile 5 | Not health adjusted | 1.21 | 1.2 | 1.21 | 0 |
| Standard acuity | IMD Decile 5 | Health adjusted | 1.14 | 1.14 | 1.15 | 0 |
| Standard acuity | IMD Decile 6 | Not health adjusted | 1.19 | 1.18 | 1.19 | 0 |
| Standard acuity | IMD Decile 6 | Health adjusted | 1.14 | 1.13 | 1.14 | 0 |
| Standard acuity | IMD Decile 7 | Not health adjusted | 1.14 | 1.13 | 1.14 | 0 |
| Standard acuity | IMD Decile 7 | Health adjusted | 1.1 | 1.1 | 1.11 | 0 |
| Standard acuity | IMD Decile 8 | Not health adjusted | 1.13 | 1.13 | 1.14 | 0 |
| Standard acuity | IMD Decile 8 | Health adjusted | 1.11 | 1.1 | 1.11 | 0 |
| Standard acuity | IMD Decile 9 | Not health adjusted | 1.09 | 1.09 | 1.1 | 0 |
| Standard acuity | IMD Decile 9 | Health adjusted | 1.07 | 1.07 | 1.08 | 0 |
| Standard acuity | NS-SEC 2 Lower managerial, administrative and professional occupations | Not health adjusted | 1.18 | 1.18 | 1.19 | 0 |
| Standard acuity | NS-SEC 2 Lower managerial, administrative and professional occupations | Health adjusted | 1.14 | 1.14 | 1.15 | 0 |
| Standard acuity | NS-SEC 3 Intermediate occupations | Not health adjusted | 1.28 | 1.28 | 1.29 | 0 |
| Standard acuity | NS-SEC 3 Intermediate occupations | Health adjusted | 1.21 | 1.2 | 1.21 | 0 |
| Standard acuity | NS-SEC 4 Small employers and own account workers | Not health adjusted | 1.37 | 1.36 | 1.38 | 0 |
| Standard acuity | NS-SEC 4 Small employers and own account workers | Health adjusted | 1.31 | 1.3 | 1.31 | 0 |
| Standard acuity | NS-SEC 5 Lower supervisory and technical occupations | Not health adjusted | 1.49 | 1.49 | 1.5 | 0 |
| Standard acuity | NS-SEC 5 Lower supervisory and technical occupations | Health adjusted | 1.37 | 1.37 | 1.38 | 0 |
| Standard acuity | NS-SEC 6 Semi-routine occupations | Not health adjusted | 1.49 | 1.48 | 1.49 | 0 |
| Standard acuity | NS-SEC 6 Semi-routine occupations | Health adjusted | 1.32 | 1.32 | 1.33 | 0 |
| Standard acuity | NS-SEC 7 Routine occupations | Not health adjusted | 1.57 | 1.56 | 1.58 | 0 |
| Standard acuity | NS-SEC 7 Routine occupations | Health adjusted | 1.4 | 1.4 | 1.41 | 0 |
| Standard acuity | NS-SEC 8 Never worked and long-term unemployed | Not health adjusted | 1.62 | 1.61 | 1.63 | 0 |
| Standard acuity | NS-SEC 8 Never worked and long-term unemployed | Health adjusted | 1.3 | 1.29 | 1.31 | 0 |
| Standard acuity | Highest qualification level 1: 1-4 GCSEs and equivalent | Not health adjusted | 1.27 | 1.27 | 1.28 | 0 |
| Standard acuity | Highest qualification level 1: 1-4 GCSEs and equivalent | Health adjusted | 1.17 | 1.16 | 1.17 | 0 |
| Standard acuity | Highest qualification level 2: 5+ GCSEs and equivalent | Not health adjusted | 1.18 | 1.17 | 1.18 | 0 |
| Standard acuity | Highest qualification level 2: 5+ GCSEs and equivalent | Health adjusted | 1.12 | 1.12 | 1.12 | 0 |
| Standard acuity | Highest qualification level 3: A levels, AS levels and equivalent | Not health adjusted | 1.17 | 1.17 | 1.17 | 0 |
| Standard acuity | Highest qualification level 3: A levels, AS levels and equivalent | Health adjusted | 1.12 | 1.12 | 1.13 | 0 |
| Standard acuity | Highest qualification: Apprenticeship | Not health adjusted | 1.3 | 1.3 | 1.31 | 0 |
| Standard acuity | Highest qualification: Apprenticeship | Health adjusted | 1.22 | 1.22 | 1.23 | 0 |
| Standard acuity | Highest qualification: no qualification | Not health adjusted | 1.37 | 1.36 | 1.37 | 0 |
| Standard acuity | Highest qualification: no qualification | Health adjusted | 1.2 | 1.2 | 1.2 | 0 |
| Urgent acuity | IMD Decile 1 | Not health adjusted | 1.64 | 1.64 | 1.65 | 0 |
| Urgent acuity | IMD Decile 1 | Health adjusted | 1.25 | 1.24 | 1.25 | 0 |
| Urgent acuity | IMD Decile 2 | Not health adjusted | 1.51 | 1.5 | 1.52 | 0 |
| Urgent acuity | IMD Decile 2 | Health adjusted | 1.21 | 1.2 | 1.22 | 0 |
| Urgent acuity | IMD Decile 3 | Not health adjusted | 1.44 | 1.43 | 1.44 | 0 |
| Urgent acuity | IMD Decile 3 | Health adjusted | 1.19 | 1.19 | 1.2 | 0 |
| Urgent acuity | IMD Decile 4 | Not health adjusted | 1.31 | 1.31 | 1.32 | 0 |
| Urgent acuity | IMD Decile 4 | Health adjusted | 1.13 | 1.12 | 1.13 | 0 |
| Urgent acuity | IMD Decile 5 | Not health adjusted | 1.22 | 1.21 | 1.22 | 0 |
| Urgent acuity | IMD Decile 5 | Health adjusted | 1.08 | 1.07 | 1.08 | 0 |
| Urgent acuity | IMD Decile 6 | Not health adjusted | 1.18 | 1.18 | 1.19 | 0 |
| Urgent acuity | IMD Decile 6 | Health adjusted | 1.08 | 1.07 | 1.08 | 0 |
| Urgent acuity | IMD Decile 7 | Not health adjusted | 1.12 | 1.12 | 1.13 | 0 |
| Urgent acuity | IMD Decile 7 | Health adjusted | 1.05 | 1.04 | 1.05 | 0 |
| Urgent acuity | IMD Decile 8 | Not health adjusted | 1.08 | 1.08 | 1.09 | 0 |
| Urgent acuity | IMD Decile 8 | Health adjusted | 1.02 | 1.02 | 1.03 | 0 |
| Urgent acuity | IMD Decile 9 | Not health adjusted | 1.04 | 1.03 | 1.04 | 0 |
| Urgent acuity | IMD Decile 9 | Health adjusted | 1 | 1 | 1.01 | 0.78 |
| Urgent acuity | NS-SEC 2 Lower managerial, administrative and professional occupations | Not health adjusted | 1.19 | 1.18 | 1.19 | 0 |
| Urgent acuity | NS-SEC 2 Lower managerial, administrative and professional occupations | Health adjusted | 1.11 | 1.11 | 1.12 | 0 |
| Urgent acuity | NS-SEC 3 Intermediate occupations | Not health adjusted | 1.3 | 1.29 | 1.31 | 0 |
| Urgent acuity | NS-SEC 3 Intermediate occupations | Health adjusted | 1.16 | 1.15 | 1.17 | 0 |
| Urgent acuity | NS-SEC 4 Small employers and own account workers | Not health adjusted | 1.37 | 1.36 | 1.37 | 0 |
| Urgent acuity | NS-SEC 4 Small employers and own account workers | Health adjusted | 1.24 | 1.24 | 1.25 | 0 |
| Urgent acuity | NS-SEC 5 Lower supervisory and technical occupations | Not health adjusted | 1.47 | 1.46 | 1.48 | 0 |
| Urgent acuity | NS-SEC 5 Lower supervisory and technical occupations | Health adjusted | 1.24 | 1.23 | 1.25 | 0 |
| Urgent acuity | NS-SEC 6 Semi-routine occupations | Not health adjusted | 1.57 | 1.56 | 1.58 | 0 |
| Urgent acuity | NS-SEC 6 Semi-routine occupations | Health adjusted | 1.26 | 1.25 | 1.27 | 0 |
| Urgent acuity | NS-SEC 7 Routine occupations | Not health adjusted | 1.6 | 1.59 | 1.61 | 0 |
| Urgent acuity | NS-SEC 7 Routine occupations | Health adjusted | 1.27 | 1.27 | 1.28 | 0 |
| Urgent acuity | NS-SEC 8 Never worked and long-term unemployed | Not health adjusted | 1.95 | 1.94 | 1.96 | 0 |
| Urgent acuity | NS-SEC 8 Never worked and long-term unemployed | Health adjusted | 1.3 | 1.29 | 1.31 | 0 |
| Urgent acuity | Highest qualification level 1: 1-4 GCSEs and equivalent | Not health adjusted | 1.33 | 1.32 | 1.33 | 0 |
| Urgent acuity | Highest qualification level 1: 1-4 GCSEs and equivalent | Health adjusted | 1.13 | 1.13 | 1.14 | 0 |
| Urgent acuity | Highest qualification level 2: 5+ GCSEs and equivalent | Not health adjusted | 1.2 | 1.2 | 1.21 | 0 |
| Urgent acuity | Highest qualification level 2: 5+ GCSEs and equivalent | Health adjusted | 1.09 | 1.09 | 1.1 | 0 |
| Urgent acuity | Highest qualification level 3: A levels, AS levels and equivalent | Not health adjusted | 1.17 | 1.17 | 1.17 | 0 |
| Urgent acuity | Highest qualification level 3: A levels, AS levels and equivalent | Health adjusted | 1.08 | 1.08 | 1.09 | 0 |
| Urgent acuity | Highest qualification: Apprenticeship | Not health adjusted | 1.31 | 1.3 | 1.31 | 0 |
| Urgent acuity | Highest qualification: Apprenticeship | Health adjusted | 1.15 | 1.14 | 1.15 | 0 |
| Urgent acuity | Highest qualification: no qualification | Not health adjusted | 1.51 | 1.51 | 1.52 | 0 |
| Urgent acuity | Highest qualification: no qualification | Health adjusted | 1.17 | 1.17 | 1.18 | 0 |
| Very urgent | IMD Decile 1 | Not health adjusted | 2.01 | 1.99 | 2.02 | 0 |
| Very urgent | IMD Decile 1 | Health adjusted | 1.4 | 1.39 | 1.41 | 0 |
| Very urgent | IMD Decile 2 | Not health adjusted | 1.73 | 1.72 | 1.75 | 0 |
| Very urgent | IMD Decile 2 | Health adjusted | 1.3 | 1.29 | 1.31 | 0 |
| Very urgent | IMD Decile 3 | Not health adjusted | 1.55 | 1.54 | 1.56 | 0 |
| Very urgent | IMD Decile 3 | Health adjusted | 1.22 | 1.21 | 1.23 | 0 |
| Very urgent | IMD Decile 4 | Not health adjusted | 1.43 | 1.42 | 1.44 | 0 |
| Very urgent | IMD Decile 4 | Health adjusted | 1.18 | 1.17 | 1.19 | 0 |
| Very urgent | IMD Decile 5 | Not health adjusted | 1.31 | 1.3 | 1.32 | 0 |
| Very urgent | IMD Decile 5 | Health adjusted | 1.13 | 1.12 | 1.14 | 0 |
| Very urgent | IMD Decile 6 | Not health adjusted | 1.28 | 1.27 | 1.3 | 0 |
| Very urgent | IMD Decile 6 | Health adjusted | 1.14 | 1.13 | 1.15 | 0 |
| Very urgent | IMD Decile 7 | Not health adjusted | 1.22 | 1.21 | 1.23 | 0 |
| Very urgent | IMD Decile 7 | Health adjusted | 1.12 | 1.11 | 1.13 | 0 |
| Very urgent | IMD Decile 8 | Not health adjusted | 1.18 | 1.17 | 1.2 | 0 |
| Very urgent | IMD Decile 8 | Health adjusted | 1.11 | 1.1 | 1.12 | 0 |
| Very urgent | IMD Decile 9 | Not health adjusted | 1.08 | 1.07 | 1.09 | 0 |
| Very urgent | IMD Decile 9 | Health adjusted | 1.03 | 1.02 | 1.04 | 0 |
| Very urgent | NS-SEC 2 Lower managerial, administrative and professional occupations | Not health adjusted | 1.21 | 1.2 | 1.22 | 0 |
| Very urgent | NS-SEC 2 Lower managerial, administrative and professional occupations | Health adjusted | 1.1 | 1.09 | 1.11 | 0 |
| Very urgent | NS-SEC 3 Intermediate occupations | Not health adjusted | 1.32 | 1.3 | 1.33 | 0 |
| Very urgent | NS-SEC 3 Intermediate occupations | Health adjusted | 1.12 | 1.11 | 1.13 | 0 |
| Very urgent | NS-SEC 4 Small employers and own account workers | Not health adjusted | 1.42 | 1.4 | 1.43 | 0 |
| Very urgent | NS-SEC 4 Small employers and own account workers | Health adjusted | 1.22 | 1.21 | 1.23 | 0 |
| Very urgent | NS-SEC 5 Lower supervisory and technical occupations | Not health adjusted | 1.51 | 1.49 | 1.52 | 0 |
| Very urgent | NS-SEC 5 Lower supervisory and technical occupations | Health adjusted | 1.19 | 1.17 | 1.2 | 0 |
| Very urgent | NS-SEC 6 Semi-routine occupations | Not health adjusted | 1.63 | 1.62 | 1.65 | 0 |
| Very urgent | NS-SEC 6 Semi-routine occupations | Health adjusted | 1.2 | 1.19 | 1.22 | 0 |
| Very urgent | NS-SEC 7 Routine occupations | Not health adjusted | 1.71 | 1.7 | 1.73 | 0 |
| Very urgent | NS-SEC 7 Routine occupations | Health adjusted | 1.24 | 1.23 | 1.25 | 0 |
| Very urgent | NS-SEC 8 Never worked and long-term unemployed | Not health adjusted | 2.17 | 2.15 | 2.19 | 0 |
| Very urgent | NS-SEC 8 Never worked and long-term unemployed | Health adjusted | 1.27 | 1.26 | 1.28 | 0 |
| Very urgent | Highest qualification level 1: 1-4 GCSEs and equivalent | Not health adjusted | 1.39 | 1.38 | 1.4 | 0 |
| Very urgent | Highest qualification level 1: 1-4 GCSEs and equivalent | Health adjusted | 1.1 | 1.09 | 1.11 | 0 |
| Very urgent | Highest qualification level 2: 5+ GCSEs and equivalent | Not health adjusted | 1.22 | 1.21 | 1.23 | 0 |
| Very urgent | Highest qualification level 2: 5+ GCSEs and equivalent | Health adjusted | 1.06 | 1.05 | 1.07 | 0 |
| Very urgent | Highest qualification level 3: A levels, AS levels and equivalent | Not health adjusted | 1.17 | 1.16 | 1.17 | 0 |
| Very urgent | Highest qualification level 3: A levels, AS levels and equivalent | Health adjusted | 1.05 | 1.04 | 1.06 | 0 |
| Very urgent | Highest qualification: Apprenticeship | Not health adjusted | 1.34 | 1.32 | 1.35 | 0 |
| Very urgent | Highest qualification: Apprenticeship | Health adjusted | 1.1 | 1.09 | 1.11 | 0 |
| Very urgent | Highest qualification: no qualification | Not health adjusted | 1.64 | 1.63 | 1.65 | 0 |
| Very urgent | Highest qualification: no qualification | Health adjusted | 1.15 | 1.14 | 1.16 | 0 |
| Immediate acuity | IMD Decile 1 | Not health adjusted | 1.22 | 1.2 | 1.25 | 0 |
| Immediate acuity | IMD Decile 1 | Health adjusted | 0.83 | 0.82 | 0.85 | 0 |
| Immediate acuity | IMD Decile 2 | Not health adjusted | 1.35 | 1.32 | 1.38 | 0 |
| Immediate acuity | IMD Decile 2 | Health adjusted | 0.99 | 0.97 | 1.01 | 0.49 |
| Immediate acuity | IMD Decile 3 | Not health adjusted | 1.46 | 1.44 | 1.49 | 0 |
| Immediate acuity | IMD Decile 3 | Health adjusted | 1.14 | 1.11 | 1.16 | 0 |
| Immediate acuity | IMD Decile 4 | Not health adjusted | 1.36 | 1.34 | 1.39 | 0 |
| Immediate acuity | IMD Decile 4 | Health adjusted | 1.11 | 1.09 | 1.13 | 0 |
| Immediate acuity | IMD Decile 5 | Not health adjusted | 1.43 | 1.4 | 1.46 | 0 |
| Immediate acuity | IMD Decile 5 | Health adjusted | 1.22 | 1.19 | 1.24 | 0 |
| Immediate acuity | IMD Decile 6 | Not health adjusted | 1.34 | 1.31 | 1.36 | 0 |
| Immediate acuity | IMD Decile 6 | Health adjusted | 1.18 | 1.16 | 1.2 | 0 |
| Immediate acuity | IMD Decile 7 | Not health adjusted | 1.32 | 1.29 | 1.35 | 0 |
| Immediate acuity | IMD Decile 7 | Health adjusted | 1.2 | 1.18 | 1.22 | 0 |
| Immediate acuity | IMD Decile 8 | Not health adjusted | 1.18 | 1.16 | 1.21 | 0 |
| Immediate acuity | IMD Decile 8 | Health adjusted | 1.1 | 1.08 | 1.12 | 0 |
| Immediate acuity | IMD Decile 9 | Not health adjusted | 1.15 | 1.12 | 1.17 | 0 |
| Immediate acuity | IMD Decile 9 | Health adjusted | 1.1 | 1.08 | 1.12 | 0 |
| Immediate acuity | NS-SEC 2 Lower managerial, administrative and professional occupations | Not health adjusted | 1.13 | 1.11 | 1.15 | 0 |
| Immediate acuity | NS-SEC 2 Lower managerial, administrative and professional occupations | Health adjusted | 1.03 | 1.01 | 1.05 | 0 |
| Immediate acuity | NS-SEC 3 Intermediate occupations | Not health adjusted | 1.27 | 1.24 | 1.29 | 0 |
| Immediate acuity | NS-SEC 3 Intermediate occupations | Health adjusted | 1.09 | 1.06 | 1.11 | 0 |
| Immediate acuity | NS-SEC 4 Small employers and own account workers | Not health adjusted | 1.42 | 1.39 | 1.45 | 0 |
| Immediate acuity | NS-SEC 4 Small employers and own account workers | Health adjusted | 1.22 | 1.2 | 1.25 | 0 |
| Immediate acuity | NS-SEC 5 Lower supervisory and technical occupations | Not health adjusted | 1.37 | 1.33 | 1.4 | 0 |
| Immediate acuity | NS-SEC 5 Lower supervisory and technical occupations | Health adjusted | 1.08 | 1.05 | 1.1 | 0 |
| Immediate acuity | NS-SEC 6 Semi-routine occupations | Not health adjusted | 1.42 | 1.39 | 1.45 | 0 |
| Immediate acuity | NS-SEC 6 Semi-routine occupations | Health adjusted | 1.07 | 1.04 | 1.09 | 0 |
| Immediate acuity | NS-SEC 7 Routine occupations | Not health adjusted | 1.49 | 1.46 | 1.52 | 0 |
| Immediate acuity | NS-SEC 7 Routine occupations | Health adjusted | 1.08 | 1.06 | 1.1 | 0 |
| Immediate acuity | NS-SEC 8 Never worked and long-term unemployed | Not health adjusted | 2.09 | 2.05 | 2.13 | 0 |
| Immediate acuity | NS-SEC 8 Never worked and long-term unemployed | Health adjusted | 1.23 | 1.2 | 1.25 | 0 |
| Immediate acuity | Highest qualification level 1: 1-4 GCSEs and equivalent | Not health adjusted | 1.33 | 1.31 | 1.35 | 0 |
| Immediate acuity | Highest qualification level 1: 1-4 GCSEs and equivalent | Health adjusted | 1.06 | 1.05 | 1.08 | 0 |
| Immediate acuity | Highest qualification level 2: 5+ GCSEs and equivalent | Not health adjusted | 1.15 | 1.13 | 1.17 | 0 |
| Immediate acuity | Highest qualification level 2: 5+ GCSEs and equivalent | Health adjusted | 1.01 | 0.99 | 1.03 | 0.37 |
| Immediate acuity | Highest qualification level 3: A levels, AS levels and equivalent | Not health adjusted | 1.07 | 1.06 | 1.09 | 0 |
| Immediate acuity | Highest qualification level 3: A levels, AS levels and equivalent | Health adjusted | 0.98 | 0.96 | 0.99 | 0.01 |
| Immediate acuity | Highest qualification: Apprenticeship | Not health adjusted | 1.21 | 1.19 | 1.24 | 0 |
| Immediate acuity | Highest qualification: Apprenticeship | Health adjusted | 1 | 0.98 | 1.02 | 0.75 |
| Immediate acuity | Highest qualification: no qualification | Not health adjusted | 1.61 | 1.59 | 1.63 | 0 |
| Immediate acuity | Highest qualification: no qualification | Health adjusted | 1.12 | 1.1 | 1.13 | 0 |
